# Combined population and patient-specific brain networks predict electrical stimulation propagation

**DOI:** 10.64898/2026.09.29.26364360

**Authors:** Sarah B. Lavelle, William K. S. Ojemann, Marc Jaskir, Zican Zou, Carlos A. Aguila, Katherine Walsh, Odile Feys, Daniel J. Zhou, Kathryn A. Davis, Brian Litt, Erin C. Conrad, Nishant Sinha

## Abstract

**Objective:** Cortico-cortical evoked potentials (CCEPs) are used during intracranial EEG (iEEG) monitoring to probe effective connectivity and guide epilepsy surgery. However, the factors governing spatial distribution and variability of stimulation-evoked responses across patients remain incompletely understood, making it difficult to distinguish physiological variation from pathological responses. We aimed to develop interpretable models of CCEP responses that integrate normative and patient-specific features, and to assess how these relationships are influenced by distance, brain-state, and epileptogenicity.

**Methods:** We analyzed single-pulse stimulation data from 34 patients with drug-resistant epilepsy undergoing stereo-EEG monitoring. Significant CCEP responses were defined using N1 peak amplitude. For each stimulation-response pair, we derived multimodal features capturing Euclidean distance, effective connectivity priors from F-TRACT, a published population atlas of CCEP response probabilities, normative structural connectivity and communication metrics, and patient-specific functional connectivity estimated from iEEG during baseline and pre-stimulation periods. Lasso-regularized logistic regression models were trained to predict response using leave-one-patient-out cross-validation, and feature importance was examined across spatial scales and seizure onset zone (SOZ) involvement.

**Results:** Performance to predict stimulation-evoked responses improved with the integration of multiple feature domains, with the full model achieving the highest accuracy (median AUC = 0.84). Feature relevance varied with spatial scale: short-range responses were dominated by distance and normative priors, whereas long-range responses depended strongly on patient-specific functional connectivity, especially when measured from pre-stimulation periods. Prediction errors were significantly higher for stimulation-response pairs within the SOZ, indicating deviations from normative connectivity-response relationships in epileptogenic networks.

**Significance:** CCEP responses reflect an interaction between spatial constraints, network architecture, and patient-specific brain state. Multimodal modeling of CCEPs provides a principled framework to interpret stimulation responses and may offer complementary markers for identifying epileptogenic networks during presurgical evaluation.

## INTRODUCTION

Direct electrical stimulation during intracranial EEG (iEEG) monitoring generates cortico-cortical evoked potentials (CCEPs), providing a causal and temporally precise assay of how activity propagates through human brain networks^1,2^. In epilepsy, CCEPs are increasingly used to probe effective connectivity^1,3^, and to test hypotheses about network excitability^4–9^, while in neuromodulation they offer a practical way to estimate downstream, off-target effects of stimulation^10–13^, an essential step as therapies move from focal targets toward network-level intervention. Despite their promise for clinical translation, the determinants of when and where CCEPs emerge across the implanted brain remain only partially understood, limiting interpretability and generalization across patients and clinical settings.

A central gap is that most mechanistic explanations of CCEP propagation emphasize a single domain (distance^14^, structural wiring^15^, or functional coupling^16^) without a unified framework that quantifies their relative contributions. Population resources such as Functional Tractography (F-TRACT) summarize conserved stimulation–response patterns across hundreds of implantations^17,18^, while normative structural connectomes capture topology and polysynaptic communication constraints^19,20^. However, it is unclear how much these population-level normative priors explain patient-level responses and whether individualized networks provide independent explanatory value beyond available effective connectivity atlases.

A second gap is that stimulation responses are known to vary with brain state (e.g. sleep vs. wakefulness)^1,21,22^, yet state-dependent predictors are rarely integrated into models that are both interpretable and generalizable. Intracranial EEG offers a unique, multiscale window into patient-specific functional coupling, and it captures at least two distinct regimes relevant to stimulation propagation. Stable baseline networks, estimated over minutes of recording, reproducible across brain states, and closely tied to underlying anatomy^23–26^, coexist with dynamic functional connectivity that fluctuates on faster timescales and more tightly tracks acute behavioral or brain-state changes^27,28^. How each regime shapes CCEP propagation, and their relative contributions, remains unresolved. Further, if normative models capture “expected” propagation, then systematic deviations from those expectations may reflect pathology^29–32^; however, the extent to which epileptogenic tissue deviates from normative evoked response patterns and relationships has not been tested in a multivariate, cross-validated framework.

Here, we build an interpretable framework to predict CCEP responses in 34 patients undergoing stereo-EEG monitoring for drug-resistant epilepsy. Specifically, we: a) model whether stimulation of a source region of interest (ROI) evokes a significant N1 response in a target ROI using five feature domains, Euclidean distance, population effective-connectivity priors (F-TRACT), normative fMRI functional connectivity, and structural connectivity/communication metrics, and patient-specific iEEG functional connectivity at baseline and immediately pre-stimulation; b) use leave-one-patient-out validation to quantify each domain’s contribution and identify consistently predictive features; and c) test two mechanistic hypotheses: that short- and long-range responses rely on distinct domains, proximal responses on conserved effective-connectivity priors and stable patient-specific functional coupling, distal responses on polysynaptic structural communication^19,33^, and that stimulation within epileptogenic networks, which are intrinsically hyperconnected yet functionally disconnected from the surrounding network^4,8^ and deviate from normative maps^29,30^, yields larger departures from model expectations, i.e. increased prediction error for seizure-onset-zone pairs.

## METHODS

### 2.1 Patient Cohort

We retrospectively analyzed data from 34 patients with drug-resistant epilepsy undergoing stereo-electroencephalography (sEEG) monitoring for epilepsy surgery planning at the Hospital of University of Pennsylvania (HUP). Stimulation sessions were performed to evoke seizures following clinical protocol and are described in detail in prior work^34,35^. All retrospective analyses present in this manuscript were approved by the HUP institutional review board (IRB protocol # 821778). Seizure onset zones (SOZ) were determined by a board-certified epileptologist from the electrographic recordings of clinically habitual seizures.

**Table 1:** Overview of Patient Cohort.

| Characteristic |  |
| --- | --- |
| Total *n* | 34 |
| Female, *n* (%) | 19 (55.8%) |
| Age at onset, years, median (range) | 22.0 (1.0-49.0) |
| Age at implant, years, median (range) | 40.5 (21.1-59.2) |
| MRI with potential epileptic lesion, *n* (%) | 12 (35.3%) |
| **SOZ lateralization [clinical determination], *n* (%)** |  |
| Right | 9 (26.5%) |
| Left | 17 (50.0%) |
| Bilateral | 5 (14.7%) |
| Inconclusive | 3 (8.8%) |
| **Intervention *n* (%)** |  |
| Resection | 9 (26.5%) |
| Laser ablation | 11 (32.3%) |
| Neuromodulation | 5 (14.7%) |
| No intervention | 9 (26.5%) |

### 2.2 CCEP acquisition protocol and data processing

Single-pulse stimulation (1 Hz, 3 mA, biphasic, 0.3-0.5 ms/phase) was delivered in 30-second trains to adjacent bipolar contact pairs; trials that evoked a stimulation-induced seizure were excluded (acquisition hardware and artifact-rejection detail in Supplementary Methods SM1). Intracranial EEG was re-referenced to a bipolar montage and segmented into stimulus-locked epochs. For each stimulation-response contact pair, trials were averaged into a mean CCEP; the N1 component was defined as the largest-magnitude peak in the 15-50ms post-stimulation window and z-scored to a pre-stimulation baseline (-500 to -30ms). A channel pair was labeled a significant response when its N1 amplitude exceeded 5 standard deviations above baseline^17^ and a non-response otherwise; this binary label was the target variable for all models.

### 2.3 Electrode Localization to ROI

Electrode contacts were localized by co-registering each patient’s post-implant CT to their pre-implant T1-weighted MRI and assigning every gray-matter contact to a ROI in the Lausanne 2008 multiscale atlas^36,37^, whose coarsest scale corresponds to the 82-region Desikan-Killiany parcellation (localization mechanics in Supplementary Methods SM2). Stimulation-response contact pairs were mapped to ROI pairs, with a single binary label per ROI pair. When multiple stimulation trials were delivered between the same pair of ROIs, a single binary label was assigned indicating whether any trial evoked a significant N1 response.

### 2.4 Multimodal Connectivity Feature Selection

For each stimulation-response ROI pair we derived features in five domains characterizing anatomical, structural, and functional connectivity used consistently throughout the analyses.

#### 2.4.1 Euclidean Distance

We computed the Euclidean distance (ED) between ROI centroids in native MRI space as a geometric proxy for anatomical separation.

#### 2.4.2 Population Effective-Connectivity Priors (F-TRACT)

We incorporated directed, frequentist cortico-cortical response probabilities from the multicenter Functional Tractography (F-TRACT) database^17,38^. These probabilities represent population-level priors on effective connectivity. To contextualize our dataset relative to this population prior, we constructed a probabilistic stimulation-response map from our cohort using the same methodology from ROI pairs sampled in more than 5 patients.

#### 2.4.3 Normative Structural Connectivity (SC)

Normative structural connectivity features were derived from running whole-brain deterministic fiber tracking on population-averaged reconstructions from the HCP1065 tractography atlas^39^. For each ROI pair, we computed connection density (CD) as the streamline count normalized by the mean volume of the two ROIs. We further derived two structural communication metrics capturing indirect, polysynaptic communication pathways^19,20^: communicability (CMY)^40^ and search information (SI)^41^.

#### 2.4.4 Normative Functional Connectivity from functional MRI (fMRI)

Normative fMRI functional connectivity features were obtained from the ENIGMA TOOLBOX^42^, Connectivity was derived from resting state fMRI from a healthy cohort in the Human Connectome Project (HCP), correlating the average blood-oxygen-level-dependent (BOLD) signal between cortical and subcortical atlas parcels and z-scoring within patients before aggregating to compute a normative functional connectome.

#### 2.4.5 Patient-Specific Functional Connectivity from intracranial EEG (iEEG)

Functional connectivity was estimated from iEEG at two timescales, a stable baseline period reflecting stable functional coupling, and a dynamic, pre-stimulation window reflecting state-specific connections, using an undirected measure (phase-locking value) and a directed measure (partial directed coherence). FC was computed on the broadband signal.

iEEG signals were bandpass filtered (1-100 Hz), notch filtered at 60 Hz and harmonics, down sampled to 256 Hz, and referenced to a common average reference across all gray matter contacts. For each stimulation-response ROI pair, two FC measures were computed: phase-locking value (PLV), an undirected measure of phase synchrony selected for robustness to interictal epileptiform activity^43^, and partial directed coherence (PDC), a directed measure that quantifies predictive information flow^4,44^.

Baseline FC was computed from a continuous 5-minute recording period preceding the stimulation session, when available. Dynamic FC was computed from a 5-second window immediately preceding each stimulation block for a given stimulation site. To compare across patients, each patient’s FC matrix was intrinsically z-scored. When multiple electrode pairs were present within a given pair of ROIs, FC values were averaged across all such electrode pairs.

### 2.5 Univariate Analyses

We performed univariate analyses to characterize marginal relationships between individual connectivity features and stimulation-evoked responses. We assessed the Pearson correlation between each feature and the binary CCEP response variable pooled across all stimulation-response ROI pairs. We further assessed partial conditional correlations correcting for Euclidean distance and F-TRACT probability for each other derived feature.

### 2.6 Multivariate Analyses

We trained logistic regression models with LASSO regularization^45^ to classify each ROI pair as evoking a significant CCEP. We then fit five nested models of increasing complexity to assess the added value of additional feature domains: ED; ED + F-TRACT; ED + F-TRACT + SC; ED + F-TRACT + SC + fMRI; ED + F-TRACT + SC + fMRI + FC. To examine feature contributions independent of population priors, secondary models excluding F-TRACT (ED + SC, ED + FC, and ED + SC + FC) were also evaluated.

#### 2.6.1 Model training and evaluation

All features were standardized z-scored prior to model training. LASSO regularization was used to perform embedded feature selection and reduce overfitting. To address class imbalance between significant and non-significant responses, we applied the synthetic minority oversampling technique (SMOTE)^46^. Model generalizability was assessed using a leave-one-patient-out cross-validation framework and performance for each held-out patient was quantified using the area under the receiver operating characteristic curve (AUC).

#### 2.6.2 Feature importance analysis

Feature importance was estimated from the absolute values of standardized model coefficients and averaged across cross-validation folds. Contributions were normalized to sum to 100% within each model, enabling comparison across feature domains. To assess scale-dependent effects, feature importance was further examined as a function of stimulation-response distance (short- vs. long-range connections) and by the presence or absence of direct structural connectivity.

### 2.7 Statistical Analysis

Confidence intervals for univariate analyses were estimated via 1,000 bootstrap resamples. Model performance across feature sets was compared using Wilcoxon signed-rank tests on patient-level AUC values obtained under leave-one-patient-out cross-validation, with Benjamini-Hochberg FDR correction across the successive-model comparisons. Feature importance (normalized absolute LASSO coefficient) is reported descriptively and stratified by distance and by structural connection. SOZ effects were tested by comparing SOZ-SOZ pairs against each other SOZ-relationship group (three comparisons): for each patient we computed the mean absolute residual and the false-positive and false-negative rates within each SOZ group, then compared the SOZ-SOZ group against each other group *within patients* using paired Wilcoxon signed-rank tests and FDR correction over the three comparisons. Clinical and epileptogenic confounders of per-patient AUC were assessed by Pearson correlation (continuous variables) and pairwise Mann-Whitney U tests with Bonferroni correction (categorical variables). Group comparisons were two-sided; tests of whether a per-patient magnitude (|ρ| or feature importance) exceeded zero were one-sided. Significance was assessed at α = 0.05. All SOZ and model analyses use the full model (every feature) and the de-duplicated all-pairs evaluation set, in which each stimulation-response pair is counted once.

## RESULTS

### 3.1 Modeling stimulation propagation with cortico-cortical evoked potentials

We performed our stimulation protocol in 34 patients, stimulating a median [IQR] of 50.5 [40.5, 66.25] channels per patient, a median of 4632.5 [2971.5, 6506.25] stimulation-response pairs. We modeled whether stimulating at one channel evoked a significant neural response elsewhere in the implanted network by thresholding the N1 cortico-cortical evoked potential (CCEP) amplitude, revealing a median of 726 [443, 889] significant response pairs per patient. After mapping stimulation-response pairs to parcellated regions to facilitate integration across patients and with normative network models, we recorded an average of 231.0 [188.0, 345.25] ipsilateral stimulation-response trials per patient with an average of 43.0 [24.25, 64.75] significant responses across stimulation sites (**Figure 1**).

**Figure 1:**
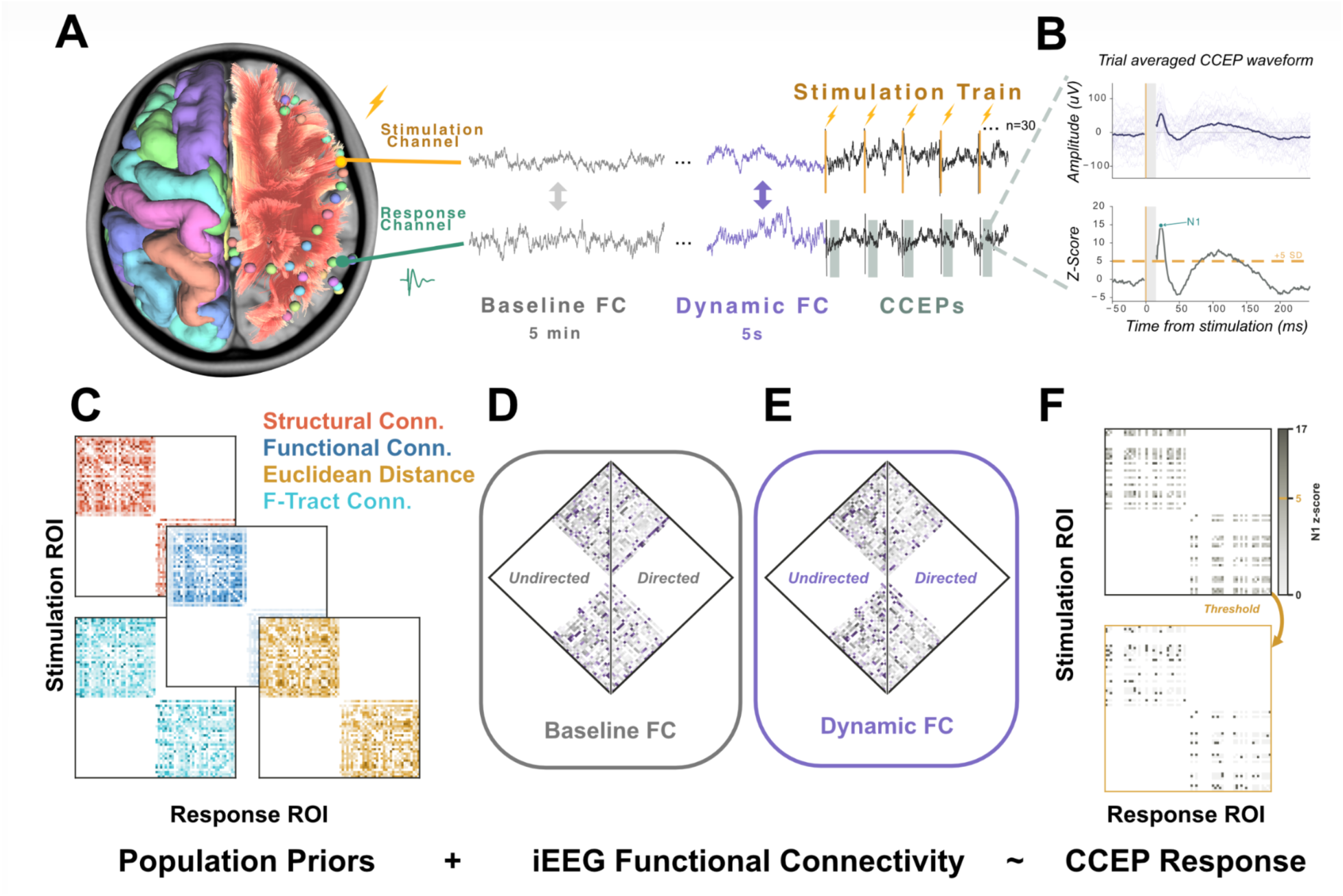
Studying the mechanisms underlying evoked responses. **(A)** Single-pulse electrical stimulation was delivered in 30-second trains to bipolar stereo-EEG contact pairs (stimulation sites) while evoked activity was recorded across the implanted network; the brain rendering shows gray matter contacts and, in the stimulated hemisphere, white matter tractography. From iEEG, functional connectivity (FC) was estimated over two timescales between the stimulating and response channels: a stable baseline period and a dynamic, state-specific period immediately preceding each stimulation block. (**B**) CCEPs were averaged over trials and normalized to baseline. Significant evoked responses were identified using a z-score threshold of 5. (**C**) Connectivity based on population priors was aggregated to estimate Euclidean distance, structural connectivity, resting-state fMRI FC, and F-TRACT population effective connectivity priors. These population priors were then combined with ROI to ROI, patient-specific FC for the baseline (**D**) and dynamic (**E**) periods, each computed with an undirected measure (phase-locking value, PLV; left half of each diamond) and a directed measure (partial directed coherence, PDC; right half). (**F**) Within a patient, every stimulated ROI pair was assessed for a significant evoked response (B), and related to the population prior and patient-specific networks.

Because stimulation and recording parameters can vary across institutions, protocols, and patients, we sought to understand the generalization of our stimulation protocol to other large, aggregated datasets of stimulation-evoked responses. At the population level, the distribution of CCEP response probabilities in our dataset strongly correlated with the independent response probabilities from the F-TRACT data consortium (R = 0.71, p <0.001; **Figure S2**). This suggests that, despite institutional variability in patient selection, implant strategy, stimulation parameters, and dataset size, a fundamental and conserved anatomical organization underlies the spatial distribution of stimulation-induced effects and motivates asking whether normative priors built on such conserved structure can explain responses in individual patients.

### 3.2 Multimodal normative brain networks constrain stimulation propagation

Given these conserved population-level patterns, we next asked which features of the aggregate brain networks constrain evoked responses. Because these predictors are population-level, we tested how well each generalizes to individual patients (**Figure 2**; **Table S1**). F-TRACT prior was the strongest correlate of individual-patient responses (|ρ| = 0.407 [0.387, 0.427]), and this association persisted even after regressing out distance (|ρ| = 0.230 [0.209, 0.251]). In contrast, and despite the documented relationship between evoked responses and BOLD connectivity^16^, we found little association between fMRI-based functional connectivity and significant CCEPs (|ρ| = 0.029 [0.008, 0.050]). Normative structural metrics from the same HCP dataset, connection density (|ρ| = 0.127 [0.101, 0.153]) and the multi-step communication measures communicability (0.163 [0.136, 0.189]) and search information (0.078 [0.058, 0.099]), were consistently, if modestly, associated with evoked potentials at the individual-patient level.

**Figure 2:**
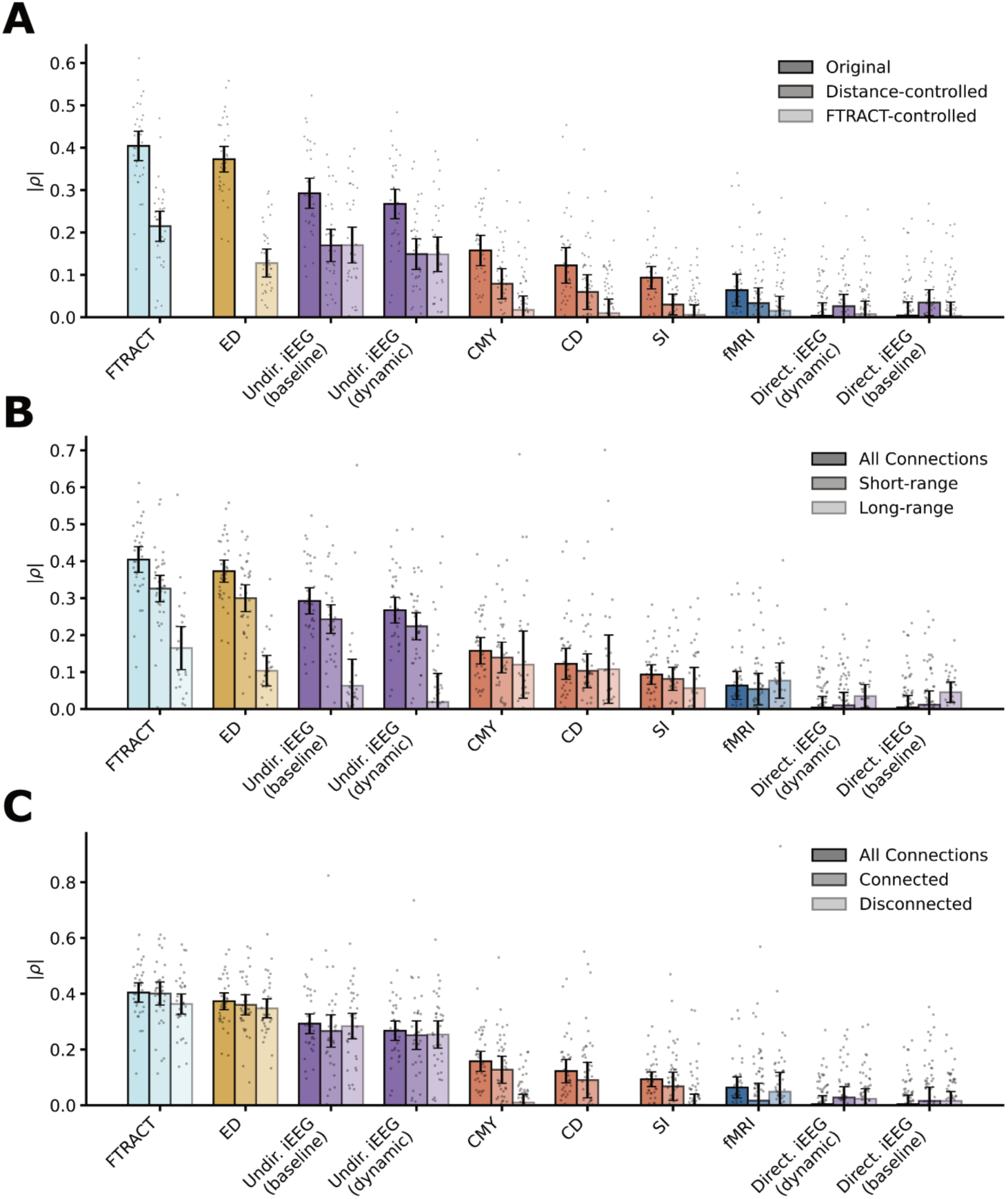
Univariate relationships between connectivity features and CCEP responses. Colors denote distance (gold), F-TRACT (turquoise), structural features (coral), fMRI (blue), and iEEG FC (purple). **(A)** Pearson correlation (|ρ|) between each connectivity feature and the binary CCEP response (significant vs. non-significant N1), computed across all stimulation–response ROI pairs pooled across patients. Features are ordered by decreasing raw correlation strength: F-TRACT response probability, Euclidean distance (ED), undirected iEEG functional connectivity (FC) at baseline, undirected iEEG FC during the dynamic (immediately pre-stimulation) period, communicability (CMY), connection density (CD), search information (SI), normative fMRI FC, and directed iEEG FC (dynamic and baseline). For each feature, correlations are shown for the original (raw) value, after regressing out Euclidean distance, and after additionally regressing out F-TRACT probability. Bars show pooled |ρ| with bootstrap 95% confidence intervals (1,000 resamples); gray points indicate per-patient correlation values. **(B)** Same analysis as in (A), stratified into short-range (<30 mm Euclidean distance) and long-range (≥30 mm) stimulation–response pairs. **(C)** Same analysis, stratified by whether stimulation–response pairs were structurally connected (nonzero white matter tract count) versus unconnected. F-TRACT probability and Euclidean distance were the strongest univariate correlates overall, and both attenuated sharply at long range (ED: 0.28 short vs. 0.08 long; F-TRACT: 0.33 vs. 0.15) but were little affected by structural connection status. Undirected iEEG FC (baseline and dynamic) was consistently associated with CCEP responses among short-range and both structurally connected/unconnected pairs (|ρ| ≈ 0.23–0.29) but was not significant at long range. The structural communication metrics (communicability, search information) were significant only among structurally connected pairs and near zero among disconnected pairs, consistent with their dependence on direct anatomical connectivity.

Some structural and imaging associations persisted, though weak, after regressing out distance (CD 0.077, CMY 0.097, SI 0.021, fMRI 0.007). However, after additionally regressing out the covariance between F-TRACT and each patient’s evoked responses, the trend vanished entirely (CD 0.023, CMY 0.026, SI 0.022, fMRI 0.011). This indicates that the structural- and BOLD-network information relevant to stimulation propagation in individual patients is already incorporated within the F-TRACT response probability (**Table S1**).

### 3.3 Patient-specific, dynamic functional networks underlie variability in CCEP response

While static networks drawn on large population priors can explain a significant portion of stimulus propagation, evoked responses have been shown to vary with rapidly changing dynamic brain state^1,6^. To assess the role of intrinsic patient-specific networks and the effect of brain-state fluctuation in predicting stimulation induced activity, we measured functional connectivity between regions implanted with iEEG electrodes during the baseline and immediately pre-stimulation periods of the same awake session (**Figure 2**; **Table S1**).

Undirected FC collected from both a baseline period (|ρ| = 0.294 [0.271, 0.317]) and directly before each new stimulation trial (|ρ| = 0.274 [0.250, 0.297]) reliably correlated with evoked responses and, unlike the normative structural metrics, these associations survived correction for both distance (baseline 0.173, dynamic 0.157) and the structural information contained in the F-TRACT prior (baseline 0.170, dynamic 0.151). In contrast, we found no reliable correlation for directed functional connectivity (baseline |ρ| = 0.000, dynamic 0.017). This suggests that while patient-specific functional connectivity, including its dynamic component, captures a brain-state signal that shapes evoked responses beyond any normative prior, directional information flow does not robustly provide the same information.

### 3.4 Multivariable models predict stimulation evoked responses

To unpack the covarying modalities, we built LASSO-regularized logistic regression models in a leave-one-patient-out cross-validation paradigm to classify stimulation-response pairs as evoking a significant N1 response (**Figure 3A**; **Table S2**). As expected, inter-regional Euclidean distance alone achieved moderate performance (median AUC 0.80 [0.76, 0.85]; AUPRC 0.44 [0.40, 0.53]). Adding normative effective-connectivity priors from F-TRACT significantly improved classification (median AUC 0.83 [0.79, 0.86]; AUPRC 0.50 [0.42, 0.58]; Wilcoxon signed-rank, FDR-corrected p < 0.001). Consistent with our univariate findings, further adding normative structural connectivity (median AUC 0.83, AUPRC 0.50) or BOLD functional connectivity (AUC 0.83, AUPRC 0.50) did not improve performance over a model combining distance and F-TRACT.

**Figure 3:**
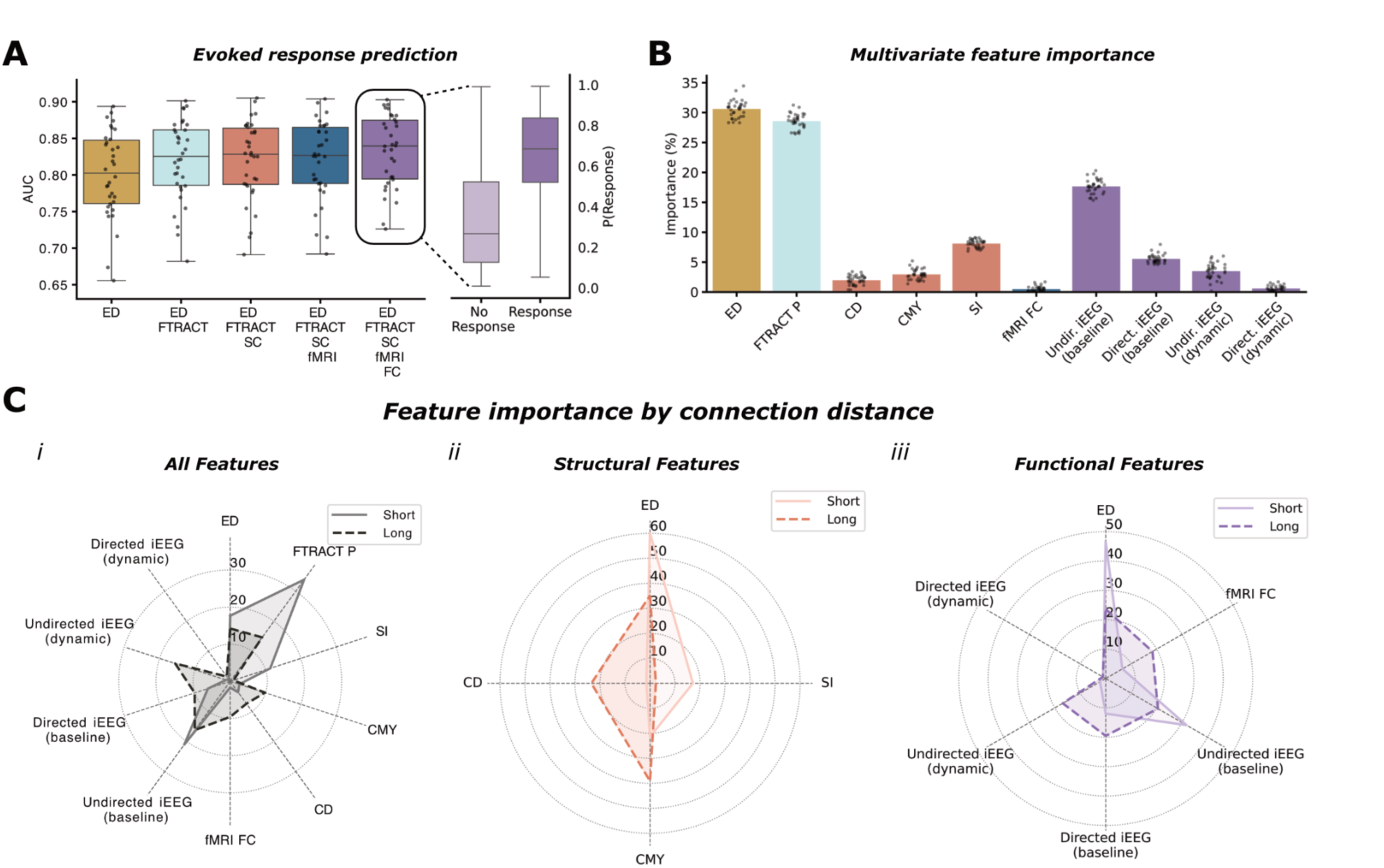
Multivariable models predict CCEP responses, with feature importance shifting by connection distance. **(A)** Performance (AUC) of nested LASSO-regularized logistic regression models predicting significant CCEP responses under leave-one-patient-out cross-validation, with feature domains added cumulatively: Euclidean distance (ED) alone; ED + F-TRACT; ED + F-TRACT + structural connectivity (SC); ED + F-TRACT + SC + fMRI; and the full model, additionally incorporating patient-specific iEEG functional connectivity (FC). Each point is one held-out patient. Right: predicted response probabilities from the full model, stratified by true CCEP outcome (no response vs. response). **(B)** Relative contribution of each feature in the full model, quantified as the mean normalized absolute LASSO coefficient (% of total model weight) averaged across cross-validation folds; points represent individual held-out patients. ED, Euclidean distance; FTRACT P, F-TRACT response probability; CD, connection density; CMY, communicability; SI, search information; fMRI FC, normative fMRI functional connectivity; Undir./Direct. iEEG, undirected/directed iEEG functional connectivity at baseline or dynamic (pre-stimulation) periods. **Colors denote distance (gold), F-TRACT (turquoise), structural features (coral), fMRI (blue), and iEEG FC (purple). (C)**Full models refit separately for short-range (<30 mm) and long-range (≥30 mm) stimulation-response pairs, shown for (i) all features, (ii) structural features only, and (iii) functional features only (radial axes rescaled per panel).

In contrast, adding patient-specific functional connectivity significantly enhanced performance. The full multimodal model (ED + F-TRACT + SC + fMRI + FC) achieved the highest overall accuracy (median AUC 0.84 [0.79, 0.87]; AUPRC 0.52 [0.43, 0.59]), significantly outperforming the preceding model (p < 0.001), and its predicted probabilities clearly separated true responses (median 0.69 [0.52, 0.84]) from non-responses (0.27 [0.13, 0.52]; **Figure 3A**).

### 3.5 Feature Importance Highlights Distinct Contributions Across Modalities

Within the full model, we quantified each feature’s relative importance as its mean normalized absolute LASSO coefficient (**Figure 3B**; **Table S3**). Results generally confirmed the findings from our univariate analyses. With the sole exception of BOLD functional connectivity (0.5%), every modality contributed to predicting CCEPs in individual patients. The two most informative features were Euclidean distance (30.6%) and the F-TRACT response probability (28.6%), together accounting for ∼59% of the total model weight. Baseline, pre-stimulation undirected FC was the third most important predictor (17.7%), underscoring the value of stable patient-specific dynamics. The remaining features each contributed less (search information 8.1%, directed baseline FC 5.5%, dynamic undirected FC 3.5%, communicability 2.9%, connection density 2.0%, dynamic directed FC 0.6%), and directed (PDC) measures were generally less predictive than their undirected counterparts. The coefficient distributions were highly consistent across held-out patients.

### 3.6 iEEG networks modulate stimulation response based on distance and anatomical connectivity

To understand how these different networks constrain evoked responses under different physical and biological conditions, we refit predictive models separately by inter-regional distance and by anatomical connection from the stimulation site (**Figure 3C**, **S3**; **Table S3**). Comparing models by distance, shorter-range responses were substantially more dependent on the conserved anatomical networks represented by F-TRACT (33.8% short vs. 14.6% long) and distance (17.7% vs. 14.2%). Conversely, the weight assigned to dynamic functional connectivity, a measure of brain state at the moment of stimulation, rose sharply with distance (15.6% long vs. 1.2% short), the largest short-to-long shift of any feature, such that undirected functional connectivity (baseline 16.2%, dynamic 15.6%) collectively dominated the long-range model. Structural weight was similar overall across scales (17.5% short vs. 18.1% long) but recomposed, with communicability and connection density rising and search information falling (11.3% short vs. 0.9% long). Splitting pairs instead by structural connection revealed a direct trade-off: distance dominated for structurally unconnected pairs (34.8% vs. 22.2% connected), whereas F-TRACT and related population priors dominated for connected pairs (31.9% vs. 22.9% unconnected; **Figure S3**). These distance- and connection-dependent shifts persisted in models refit without F-TRACT as a covariate, indicating they are independent of F-TRACT covariance.

### 3.7 Normative models of stimulation propagation encode pathological abnormalities

Because the models are trained on predominantly normative tissue, they constitute a normative model of expected responses; we therefore asked whether deviations would emerge in pathological, hyperexcitable tissue (**Figure 4**; **Table S4**).

**Figure 4:**
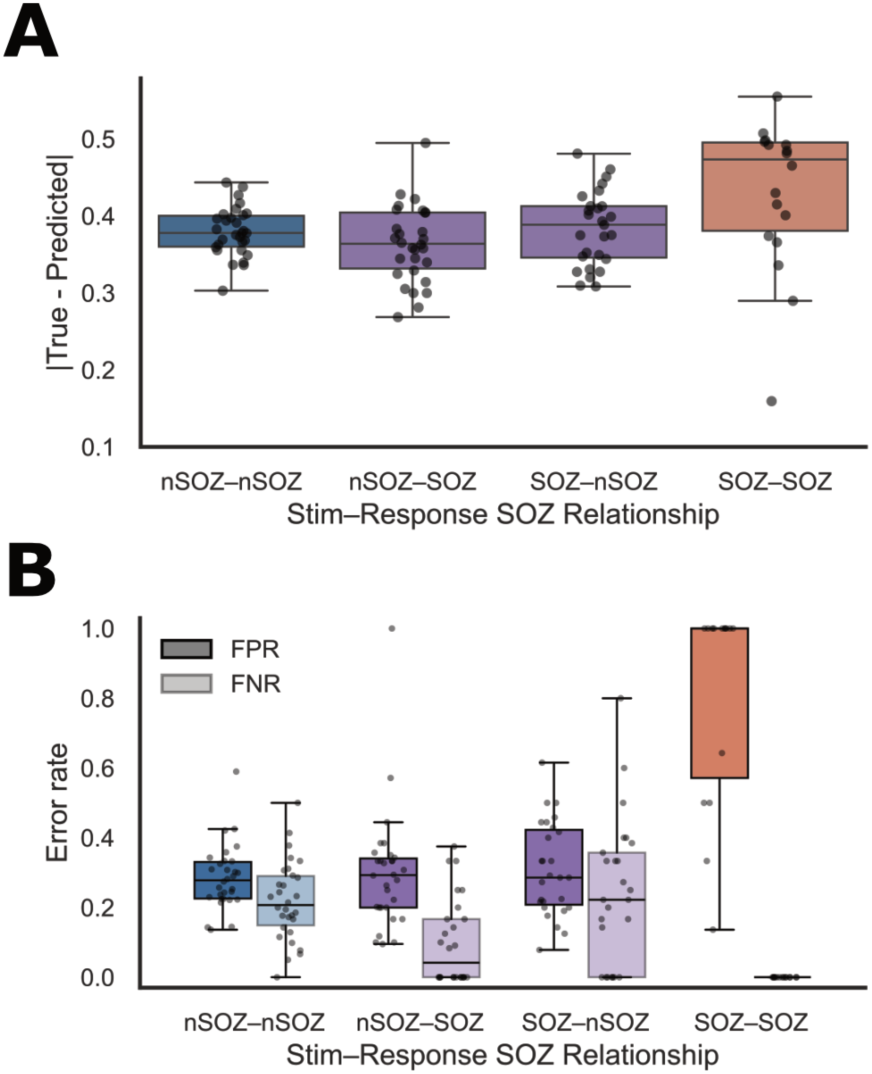
Normative models of stimulation propagation reveal pathological deviations within the seizure onset zone (SOZ). **(A)** Absolute prediction residuals (|true - predicted|) from the full multivariate model, stratified by the SOZ membership of the stimulating and responding regions: neither region in the SOZ (nSOZ-nSOZ), responding region only (nSOZ-SOZ), stimulating region only (SOZ-nSOZ), or both regions within the SOZ (SOZ-SOZ). Each point represents one patient’s mean residual within a given SOZ group. **(B)** False-positive rate (FPR, dark) and false-negative rate (FNR, light) of the full model’s predictions, stratified by the same four stimulation-response SOZ relationships. Each point represents one patient. Prediction residuals were highest for SOZ-SOZ pairs (median 0.48 vs. 0.35-0.39 for other groups), driven by a near-categorical over-prediction bias: FPR for SOZ-SOZ pairs was far higher (median 1.00) than any other group (0.28-0.31), while FNR was correspondingly near zero (median 0.00 vs. 0.04-0.22).

Consistent with this hypothesis, the clearest deviation was a systematic over-prediction of responses (predicting a significant N1 response when none met the detection criterion) when both the stimulating and recording regions fell within the seizure onset zone (SOZ-SOZ). Across patients, the model’s false-positive rate for SOZ-SOZ pairs was far higher (median 1.00) than for any other SOZ relationship (≈0.28-0.31), while its false-negative rate was correspondingly near zero (median 0.00 vs. 0.04-0.22; Figure 4B). Because most patients contribute pairs to several SOZ groups, each patient’s rate in the SOZ-SOZ group was compared against that same patient’s rate in each other group (within-patient paired Wilcoxon signed-rank, SOZ-SOZ vs. each, FDR-corrected over three comparisons; n = 15 patients with SOZ-SOZ pairs): every comparison was significant for both the false-positive rate (all p ≈ 0.001, r ≈ 0.87) and the false-negative rate (all p ≤ 0.01, r ≥ 0.88; Table S4). In effect, the model expects a response for nearly every SOZ-SOZ pair.

This over-prediction was mirrored in the prediction residuals (|true - predicted|), which were highest for SOZ-SOZ pairs (median 0.48 [0.33, 0.50]) and significantly exceeded both single-SOZ conditions (vs. nSOZ-SOZ p = 0.037, r = 0.53; vs. SOZ-nSOZ p = 0.037, r = 0.56; paired Wilcoxon, n = 18); the difference from the fully normative nSOZ-nSOZ baseline (median 0.35) was a non-significant trend (p = 0.071, r = 0.43), and pairs with only one region in the SOZ did not differ from baseline. Deviations from normative predictions were thus most robust, and directional, when both regions participate in the epileptogenic network—consistent with the model capturing the elevated hypersynchrony of the SOZ as an inflated expected response.

## DISCUSSION

Brain stimulation is central to probing and modulating distributed neural networks in epilepsy^1^ and is increasingly used for treatment planning^5,13,47^, yet the determinants of where stimulation effects emerge across the implanted brain remain only partly understood^14,19^. Here we built interpretable models of cortico-cortical evoked potentials (CCEPs) that integrate spatial, population effective-connectivity, normative structural, and patient-specific functional features. We used leave-one-patient-out validation to separate the conserved determinants of stimulation propagation from patient-and state-specific ones. CCEP detections were reviewed by a board-certified epileptologist, and data were retained using a positive predictive value (PPV) threshold >0.7.

Our central positive result is that patient-specific iEEG functional connectivity explains variance beyond population priors. Prior work established that structural, functional, and spatial factors together predict cortex-wide stimulation effects, but did so at the group level, predicting aggregate population response probabilities^19^. We asked the complementary question: given that prior, what remains to be explained in an individual patient? Undirected functional connectivity improved held-out prediction over distance, population priors, and normative imaging combined. Among these patient-specific measures, connectivity estimated from stable baseline recordings was more informative than the immediately pre-stimulation window. These findings are consistent with intracranial networks containing a persistent scaffold that emerges over minutes and is reproducible across cognitive states^23,26^. The more tractable clinical measurement is therefore also the more useful one.

At a high level, our results demonstrate that CCEP responses reflect an interplay between spatial proximity, conserved effective connectivity, and dynamic brain state. Distance and the population prior were the two strongest predictors, consistent with conduction distance and conserved cortico-cortical pathways setting the baseline likelihood of a response^14,17^. They are not independent: population response probability is itself strongly distance-dependent by construction^17^. The redundancy was asymmetric: the prior retained its association after adjustment for distance while distance largely lost its own after adjustment for the prior. This suggests that conserved anatomo-functional organization, rather than geometry alone, is the operative constraint. Importantly, however, distance and population priors together were insufficient to explain patient-specific variability, motivating the inclusion of individualized functional measures.

A key contribution of this work is the demonstration that patient-specific iEEG functional connectivity provides explanatory value beyond normative priors. Earlier work established that structural, functional, and spatial factors together predict cortex-wide stimulation effects, but did so at the group level, predicting aggregate population response probabilities^19^; we asked the complementary question of what remains to be explained within an individual patient. Undirected functional connectivity, particularly when estimated from longer stable recordings, was the most informative individualized feature, suggesting that relatively stable LFP coherence networks, which emerge over minutes and are reproducible across cognitive states^23,25,26^, strongly modulate stimulation propagation. Directed measures contributed less consistently, which we read as a dissociation rather than a measurement failure, since directed measures distinguish epileptogenic tissue^4,44^ and therapeutic neurostimulation targets^5,10,11^: the mechanisms gating whether a pulse evokes a response appear distinct from those producing spontaneous directional flow. Notably, we observed no independent contribution from imaging-based functional connectivity once the F-TRACT prior was included. This does not contradict evidence that intrinsic functional architecture predicts evoked responses^16^: that earlier study used each patient’s own resting-state fMRI rather than a population atlas; rather, population effective connectivity is itself well explained by structural, functional, and spatial factors^19^, so it largely subsumes the imaging-relevant information. While patient-specific imaging could enhance fully individualized stimulation models, our results suggest population-derived effective connectivity captures much of what is needed to predict CCEP propagation, limiting the incremental benefit of additional normative imaging features in this framework.

Our analyses further reveal that the relevance of different feature domains depends on spatial scale. Short-range pairs were dominated by distance and conserved priors, whereas patient-specific functional connectivity carried the greatest weight at long range, with its state-dependent component increasing most steeply with distance. This local-to-distal reorganization is consistent with contact-level work showing that resting functional connectivity best matches evoked responses near the stimulation site whereas structural connectivity best matches distant responses^33^, and with accounts in which long-range interactions are more sensitive to global network topology^19^. From a neuromodulation perspective, it suggests that local network engagement may be achievable with relatively static stimulation paradigms, whereas targeting distributed networks may require monitoring or leveraging state-dependent connectivity.

Beyond prediction, our framework provides insight into epileptogenic network pathology. Because the models fit predominantly on non-epileptogenic tissue, they constitute a normative expectation for the mapping of evoked response, extending normative iEEG mapping^29,30^, to abnormality in the relationship between connectivity and stimulus responsiveness. Stimulation-response pairs within the SOZ showed elevated prediction error, and the effect was most pronounced when both stimulating and responding regions lay within the SOZ, where the model systematically over-predicted responses. One possible explanation is a decoupling between intrinsic connectivity and evoked responsiveness, consistent with reports that SOZ tissue is intrinsically hyperconnected yet functionally disconnected from the surrounding network^4,8^. A model associating elevated intrinsic connectivity with greater responsiveness could then over-predict SOZ responses. These deviations highlight the potential utility of CCEP modeling not only for predicting stimulation effects but as a complementary marker of pathological network organization during presurgical evaluation. SOZ-SOZ overprediction may partly reflect spatial proximity; distance-matched non-SOZ stimulation-response pairs would help evaluate this possibility.

From a clinical perspective, our findings have two implications. First, because these relationships generalized across held-out patients and performance was independent of clinical, implant, and epileptogenic-network characteristics, multimodal modeling may reduce the need for exhaustive empirical CCEP mapping by enabling probabilistic estimation of downstream stimulation effects in new patients. This matters as therapy moves from single nodes toward networks: such models could support planning that aims to engage (or avoid) specific pathways, and connectivity-matched targets have been shown to suppress epileptiform activity where unmatched targets do not^13^, while CCEP-derived connectivity already informs electrode placement and outcome^5,12^. Second, systematic deviations from model predictions within epileptogenic networks may provide additional information for localizing seizure-generating circuits and refining surgical targets.

Our study has several limitations. We binarized CCEP responses on the presence of a significant N1 component, prioritizing robustness and interpretability over graded amplitude and latency information, which can be partly distinct^19^. We computed connectivity on the broadband signal, whereas the persistent intracranial network template is frequency-band dependent^23^, and the iEEG-BOLD correspondence is specific to high-frequency broadband power^26^. Our dynamic window sampled seconds within a single awake session, probing within-state fluctuation rather than the arousal-state transitions across which evoked responses change substantially^21,22,27,28^, which may explain why it added less than the stable baseline. Synthetic minority oversampling^46^ shifts predicted probabilities toward the response class, so we cannot fully exclude that it accentuated the SOZ over-prediction. Because population priors derive from CCEPs in drug-resistant epilepsy without explicit modeling of seizure-onset zones^17^, our prior is not a true healthy-brain normative reference, although residual epileptogenic signal should bias toward the null. Finally, although our cohort is comparable to many intracranial stimulation studies, it remains modest relative to the dimensionality of the network features examined, and larger multi-center datasets will be important for validation; we relied on normative structural connectomes rather than fully individualized diffusion imaging, though supplementary analyses suggest this did not materially affect our conclusions; seizure onset zones were defined clinically rather than by postsurgical outcome; and we applied linear models to preserve interpretability, where nonlinear approaches may better capture higher-order interactions. The risk of overfitting remains a limitation despite LASSO regularization and leave-one-patient-out validation.

In summary, our results provide mechanistic insight into how CCEPs arise from the interaction of spatial constraints, conserved network architecture, and patient-specific functional dynamics. While normative priors offer strong predictive utility, individualized brain states play a critical role in shaping stimulation-induced responses, and epileptogenic networks introduce systematic, directional deviations from expected propagation patterns. Multimodal CCEP modeling thus represents a principled framework for both guiding stimulation-based interventions and probing network integrity in epilepsy.

## Data Availability

The intracranial EEG data used in this study are available through iEEG.org (https://www.ieeg.org).

## SUPPLEMENTAL MATERIAL

### SUPPLEMENTARY METHODS

#### SM1. CCEP acquisition hardware and artifact rejection

Depth electrodes had 6–12 cylindrical contacts (2.41 mm length, 1.1 mm diameter, 5 mm inter-contact spacing). Single-pulse stimulation was delivered with a Nicolet Cortical Stimulator (Natus) across all or a subset of adjacent bipolar pairs. iEEG was sampled at 512-1024 Hz. Trials with extreme voltages (> 1000 µV) were excluded, and additional artifact rejection was applied to the averaged waveforms. CCEP detections were reviewed by a board-certified epileptologist, and data were retained using a positive predictive value (PPV) threshold >0.7. Channels with high impedance, outside the brain, or in dural proximity were excluded from stimulation, and trials producing a stimulation-induced seizure were removed^34^.

#### SM2. Electrode localization mechanics

Contacts were localized with iEEG-recon^48^: post-implant CT was rigidly registered (6 DOF) to pre-implant T1 MRI, contacts were semi-automatically identified in VoxTool, and each contact was assigned to the Lausanne 2008 (scale 33; 82 regions) ROI of highest volumetric overlap with a 2 mm-radius sphere centered at the contact. Only gray-matter contacts were retained. A pair was labeled responsive if any trial between its two ROIs evoked a significant N1.

#### SM3. Quantifying SOZ extent (number of SOZ electrodes)

From the clinically determined seizure-onset-zone electrode list for each patient, we computed the number of SOZ electrodes, setting patients with no recorded seizures to zero and excluding patients without an SOZ label (n = 33 with usable labels). This count indexes the spatial extent of the epileptogenic zone and is used in Figure S5 (its distribution by SOZ and implant laterality) and Figure S6 (against patient AUC).

## SUPPLEMENTARY RESULTS

### SR1. Model predictions are robust to clinical variation *(Figures S4, S5; Table S5)*

The relationships between population and patient-specific networks described by our models have translational potential for guiding stimulation therapy, which would require performance that is robust across the heterogeneity of clinical practice. We therefore tested whether patient-level model performance could be explained by clinical or implant-strategy variables. Although performance varied across patients (full-model AUC 0.84 [0.79, 0.87]), it was not associated with any continuous covariate (Figure S4), mean Euclidean distance (r = 0.13, p = 0.48), number of stimulation-response pairs (r = -0.15, p = 0.40), connection rate (r = 0.02, p = 0.93), response rate (r = -0.26, p = 0.14), or long- and short-range pair rates (|r| = 0.19, p = 0.28), indicating that performance is not driven by implant density or sampling. Likewise, AUC did not differ by sex (p = 0.92), surgical intervention (all Bonferroni p = 1.0), implant laterality (all Bonferroni p > 0.6), or imaging findings on MRI (potential epileptogenic lesion vs. nonspecific abnormality vs. normal; all Bonferroni p = 1.0; Figure S5). Model performance is therefore stable across the clinical variation of the cohort, supporting its potential generalization to new patients.

### SR2. Performance is not associated with SOZ lateralization or extent *(Figures S6, S7; Table S5)*

We next tested whether performance varied with the epileptogenic network itself, using the curated SOZ labels now available for 30 of the 34 patients (Figure S5). No epileptogenic-network variable was significantly associated with AUC after correction. Right-lateralized SOZs had numerically higher AUC than left-lateralized SOZs (median 0.87 [0.86, 0.88] vs. 0.83 [0.78, 0.86]), but this difference did not survive multiple-comparison correction (Mann-Whitney, raw p = 0.032, Bonferroni p = 0.097), and neither bilateral SOZs nor SOZ region/localization differed (all Bonferroni p > 0.2). Performance was likewise independent of the *extent* of the epileptogenic zone: the number of clinically determined SOZ electrodes did not correlate with patient AUC (r = -0.03, p = 0.88, n = 33; Figure S6). Descriptively, SOZ-electrode counts tracked laterality and implant breadth rather than performance—patients with bilateral SOZs carried more SOZ electrodes (median 12.5) than left- (5.5) or right-lateralized (7.0) patients, and contralateral-skewed implants more than unilateral ones (Figure S5). Together with SR1, model performance was not explained by any clinical, implant, or epileptogenic-network characteristic of the cohort. (The cohort still contains more left- than right-lateralized SOZs, 16 vs. 8, consistent with greater clinical detection of left-hemisphere seizures; the residual numerical right > left trend should be read in that light.)

## SUPPLEMENTARY FIGURES

**Figure S1 :**
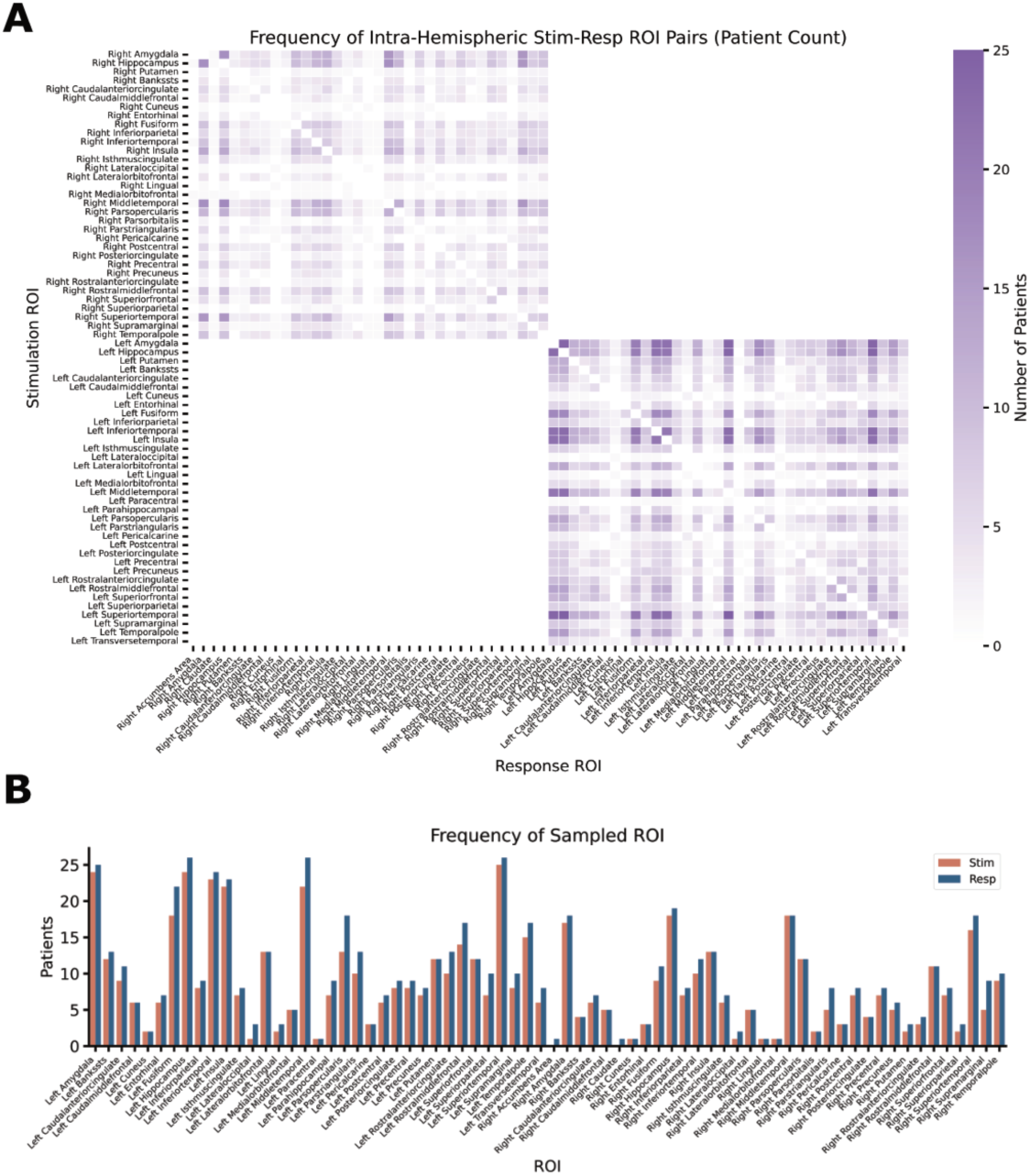
Frequency of Intra-Hemispheric stimulation-response ROI pairs in the HUP dataset. **(A)** The number of samples within a specific stimulation and response pair across the HUP dataset at the ROI level according to DK atlas. The rows are stimulation ROIs, the columns are response ROIs and the elements are the number of patients. **(B)** Number of patients with samples in specific ROIs, stratified by stimulation and response.

**Figure S2 :**
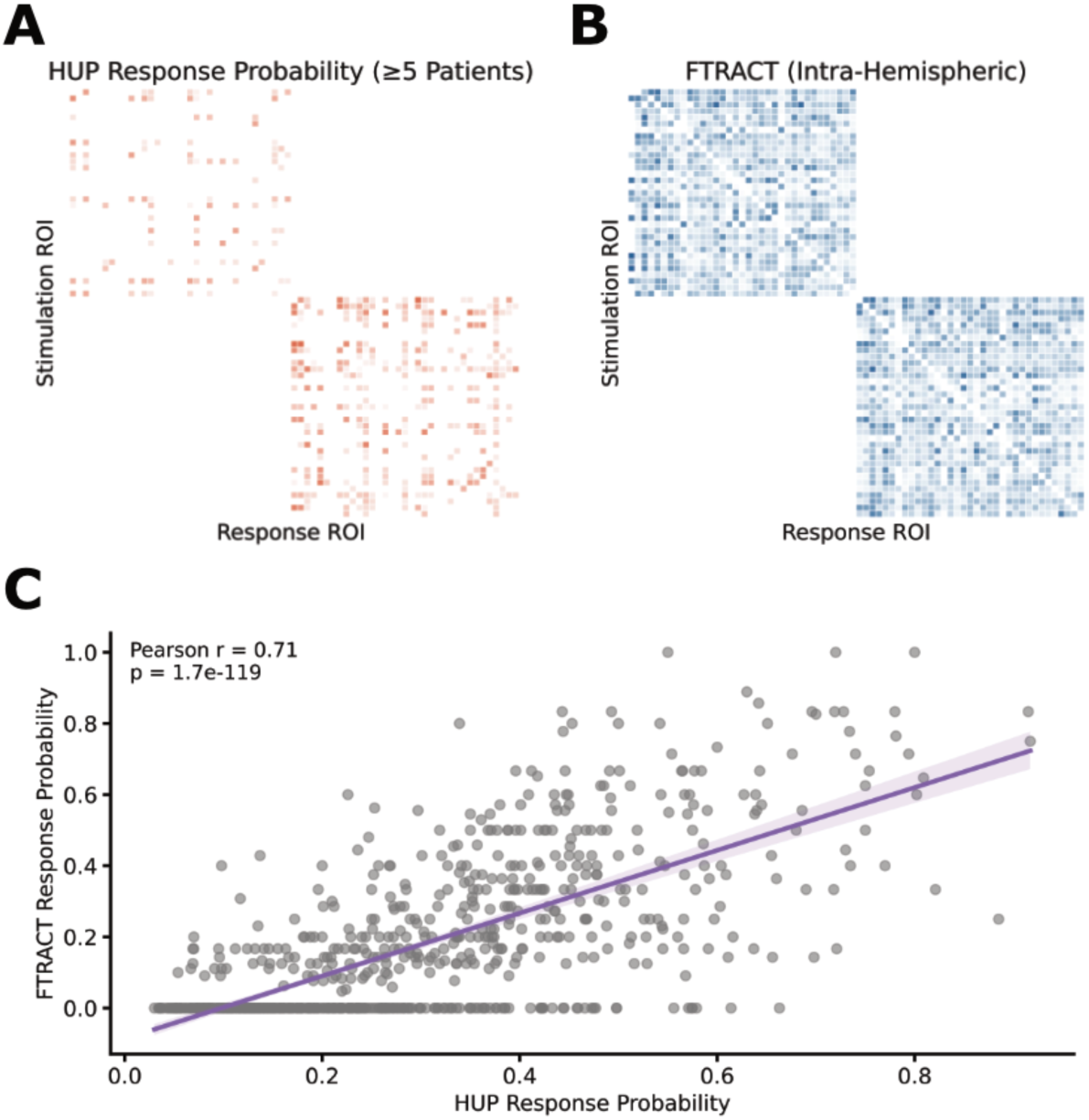
Correlation between F-TRACT response probabilities and HUP response probabilities.

**Figure S3:**
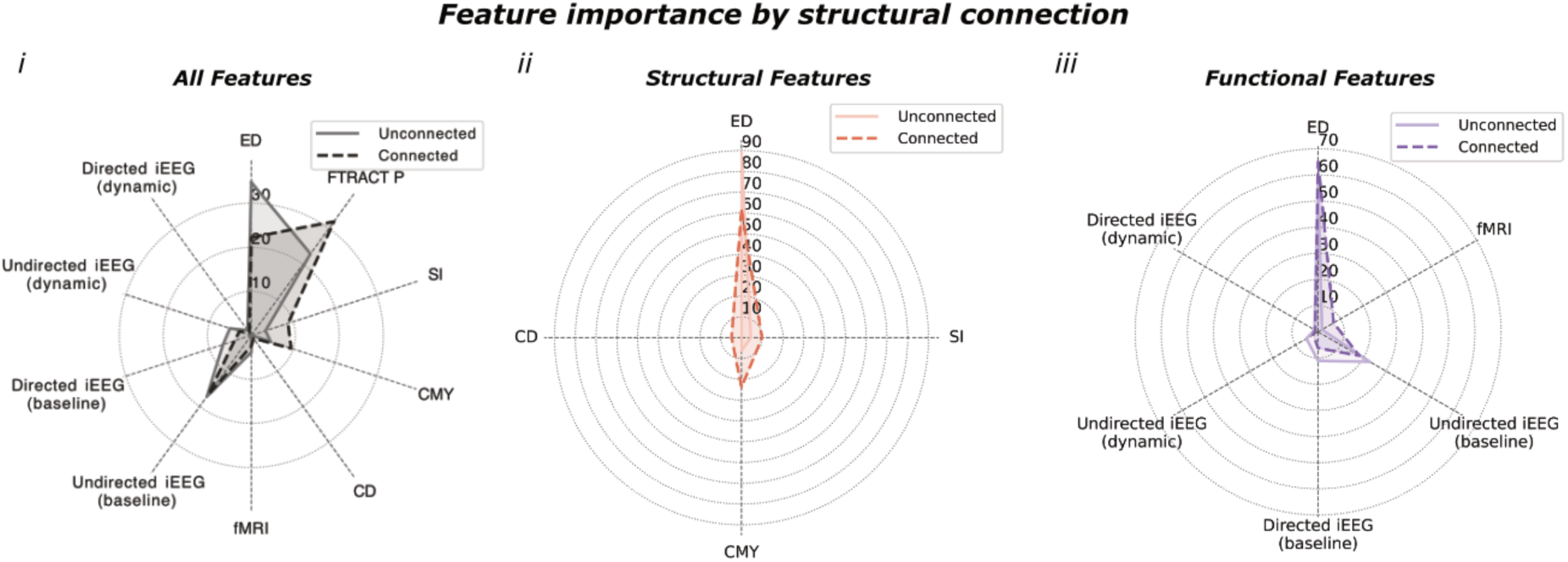
Comparison of feature importance between model with structurally connected vs unconnected stimulation-response pairs.

**Figure S4:**
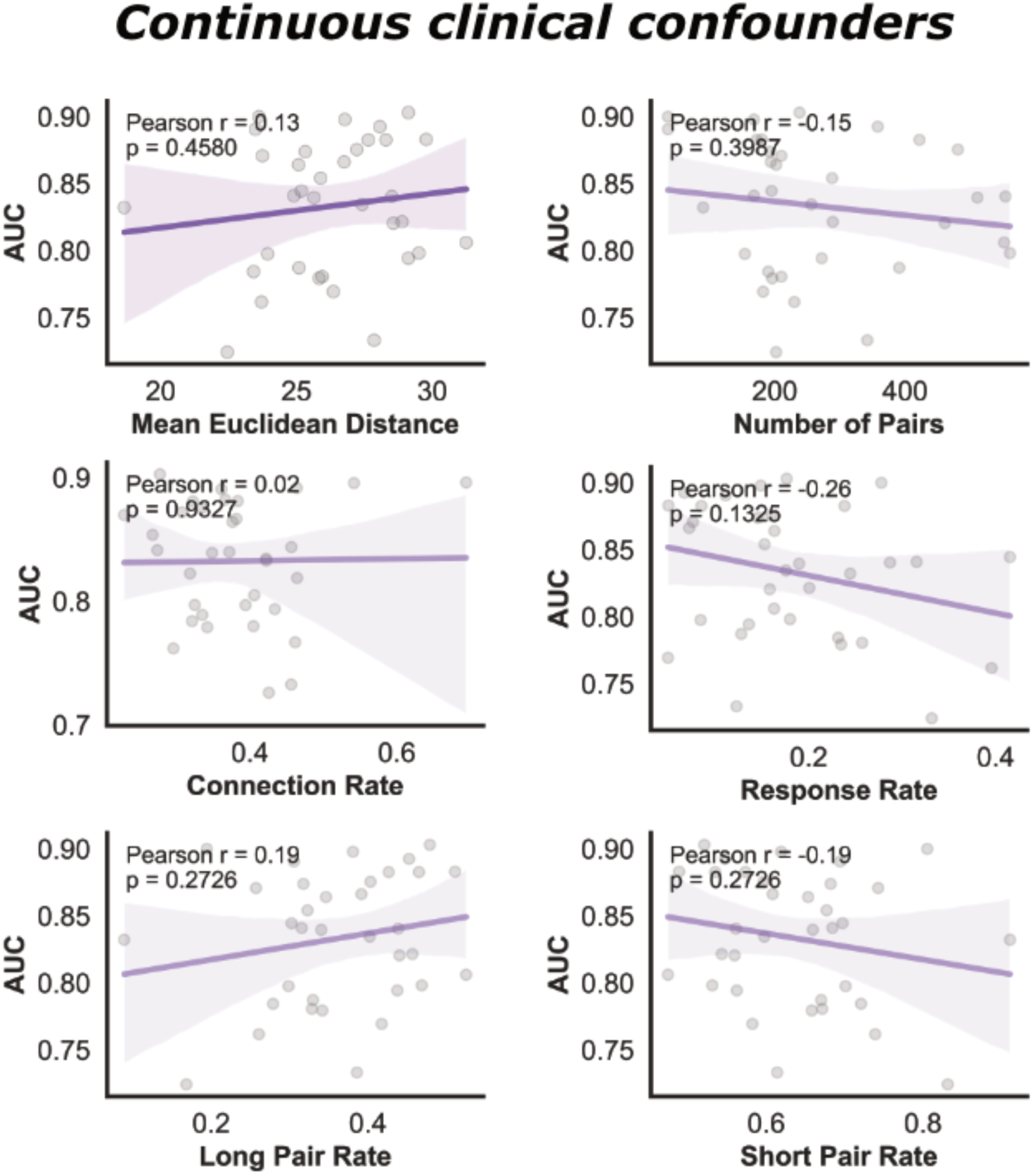
Correlation between patient-level AUCs and continuous clinical confounders.

**Figure S5:**
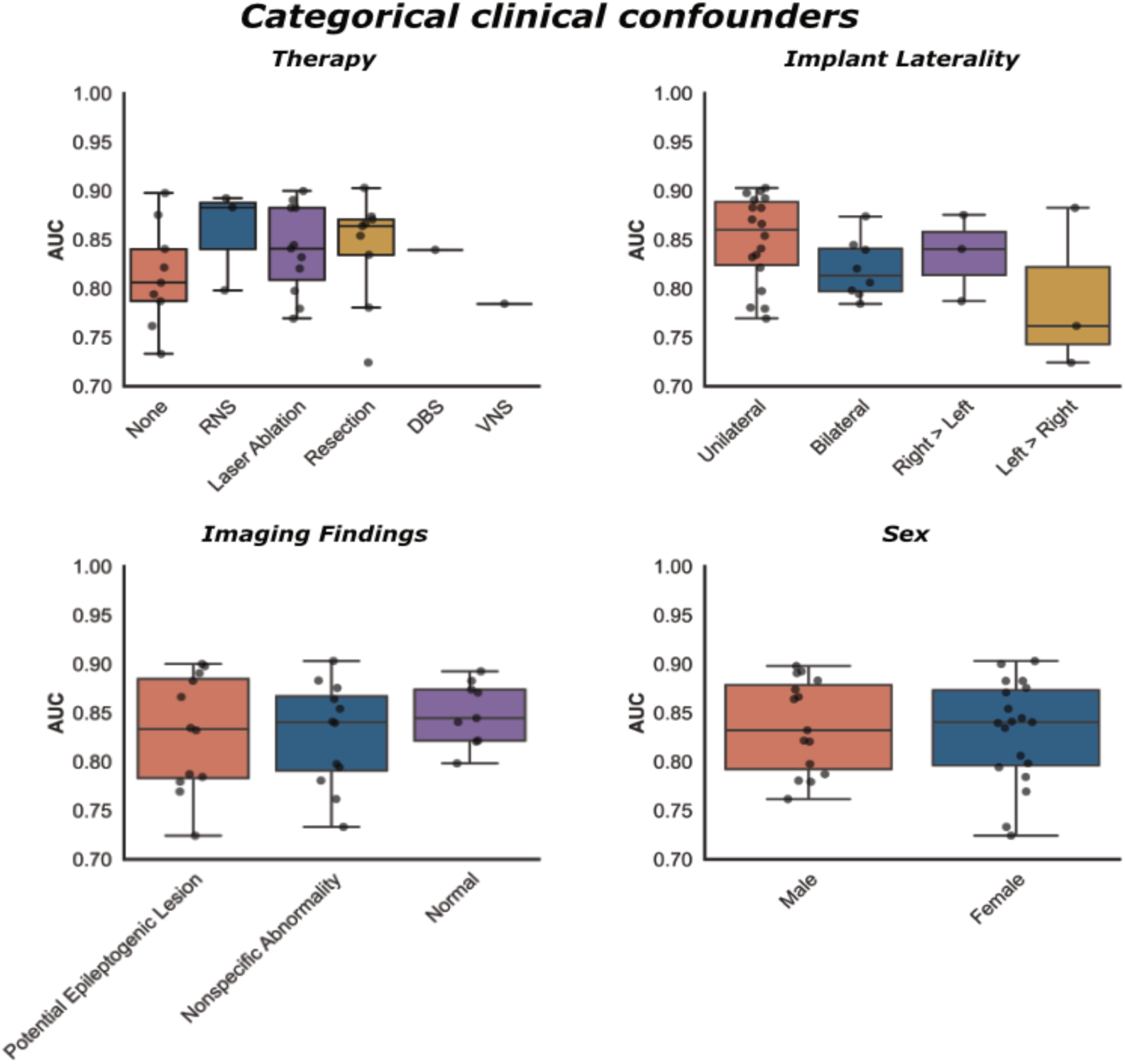
Comparison between patient-level AUCs and categorical clinical confounders.

**Figure S6:**
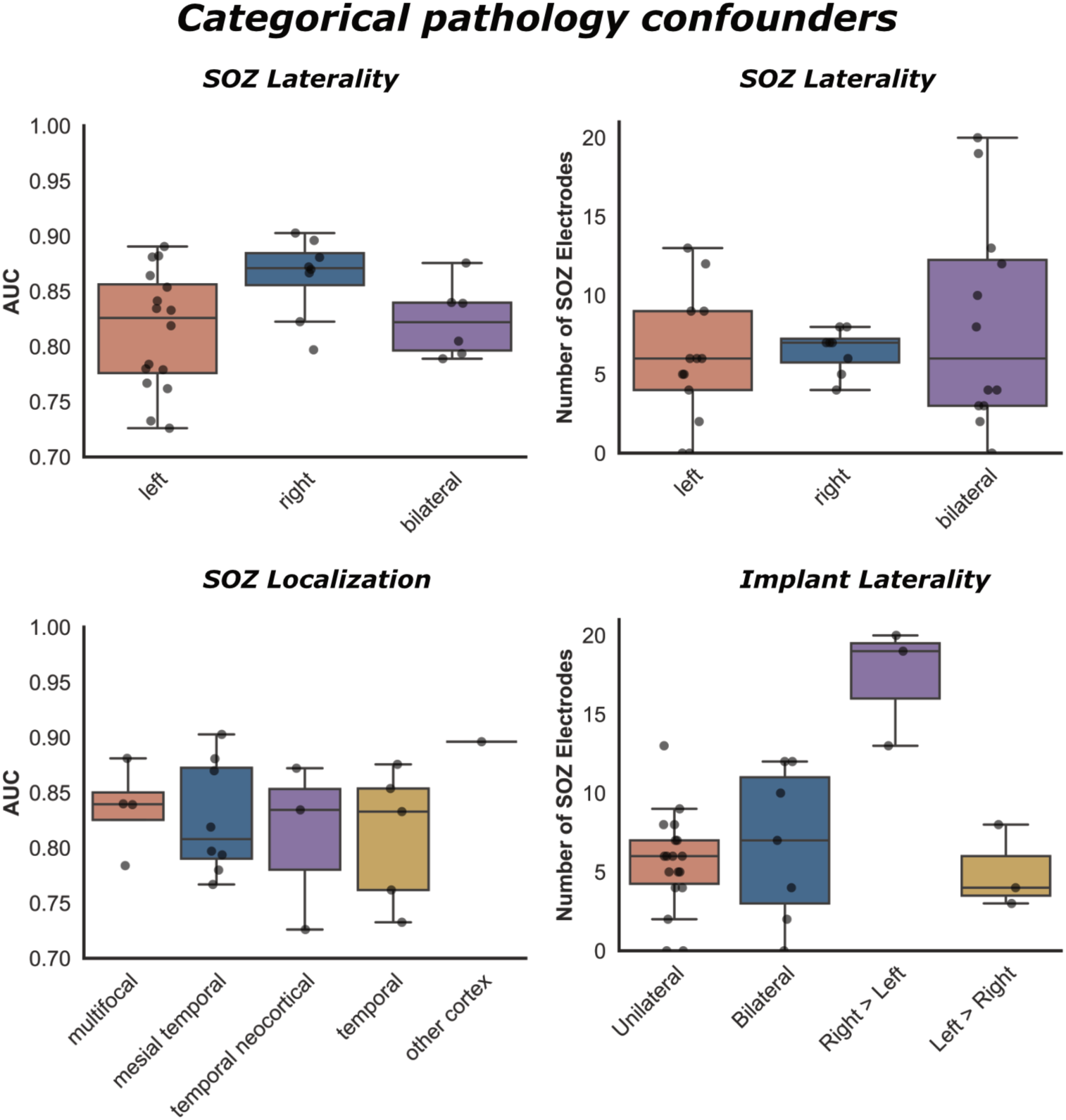
Comparison between patient-level AUCs and categorical pathology confounders.

## SUPPLEMENTARY TABLES

**Table S1 :**
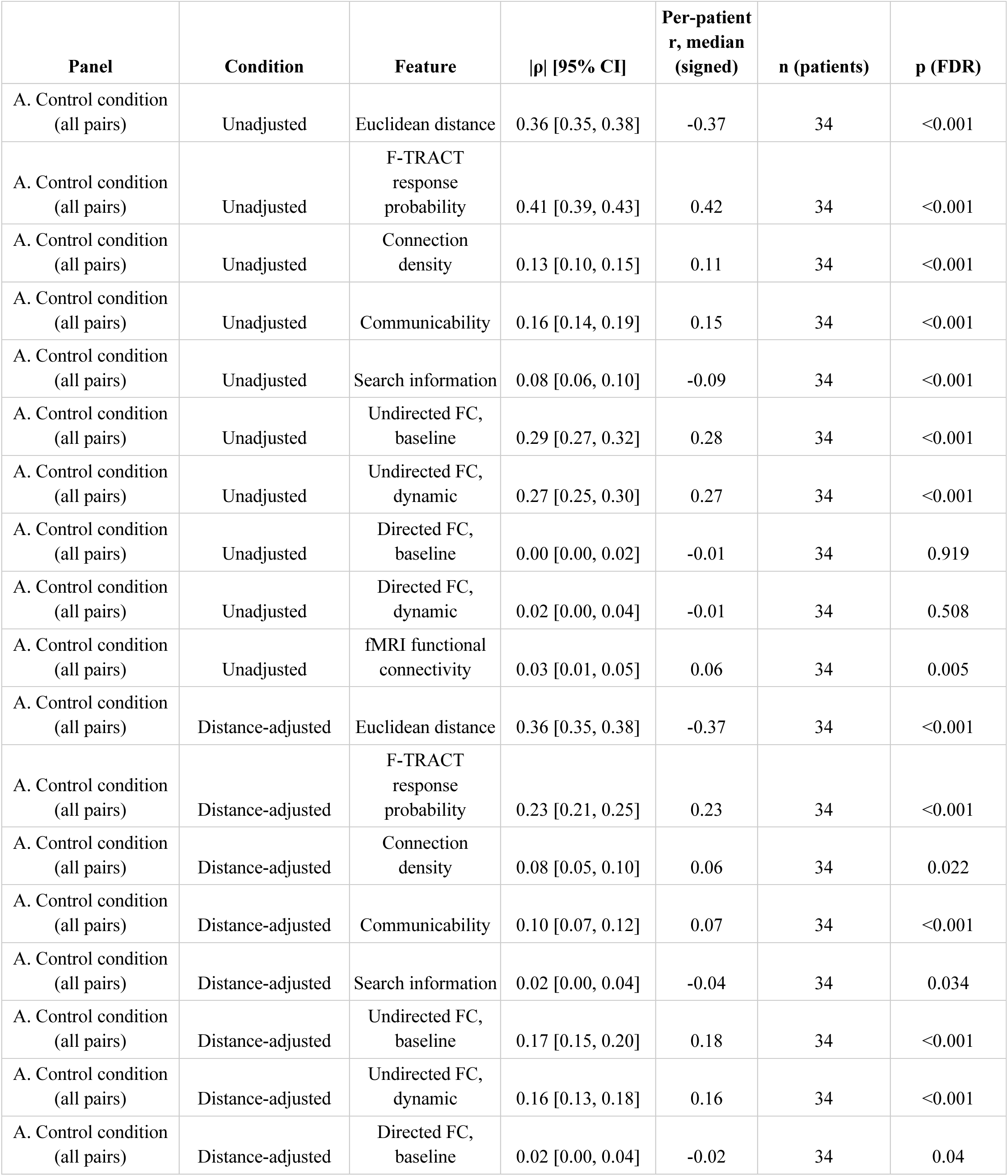
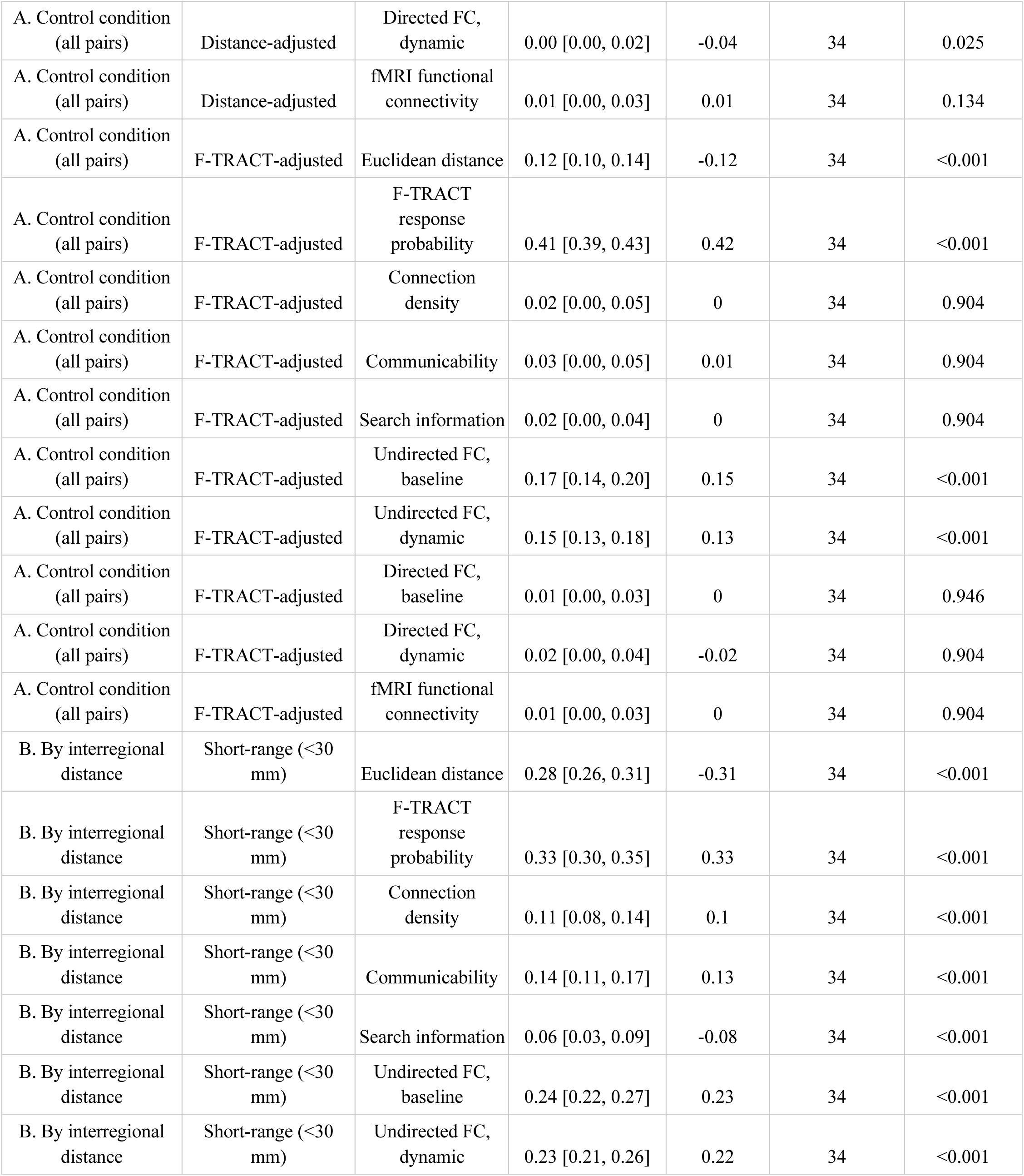
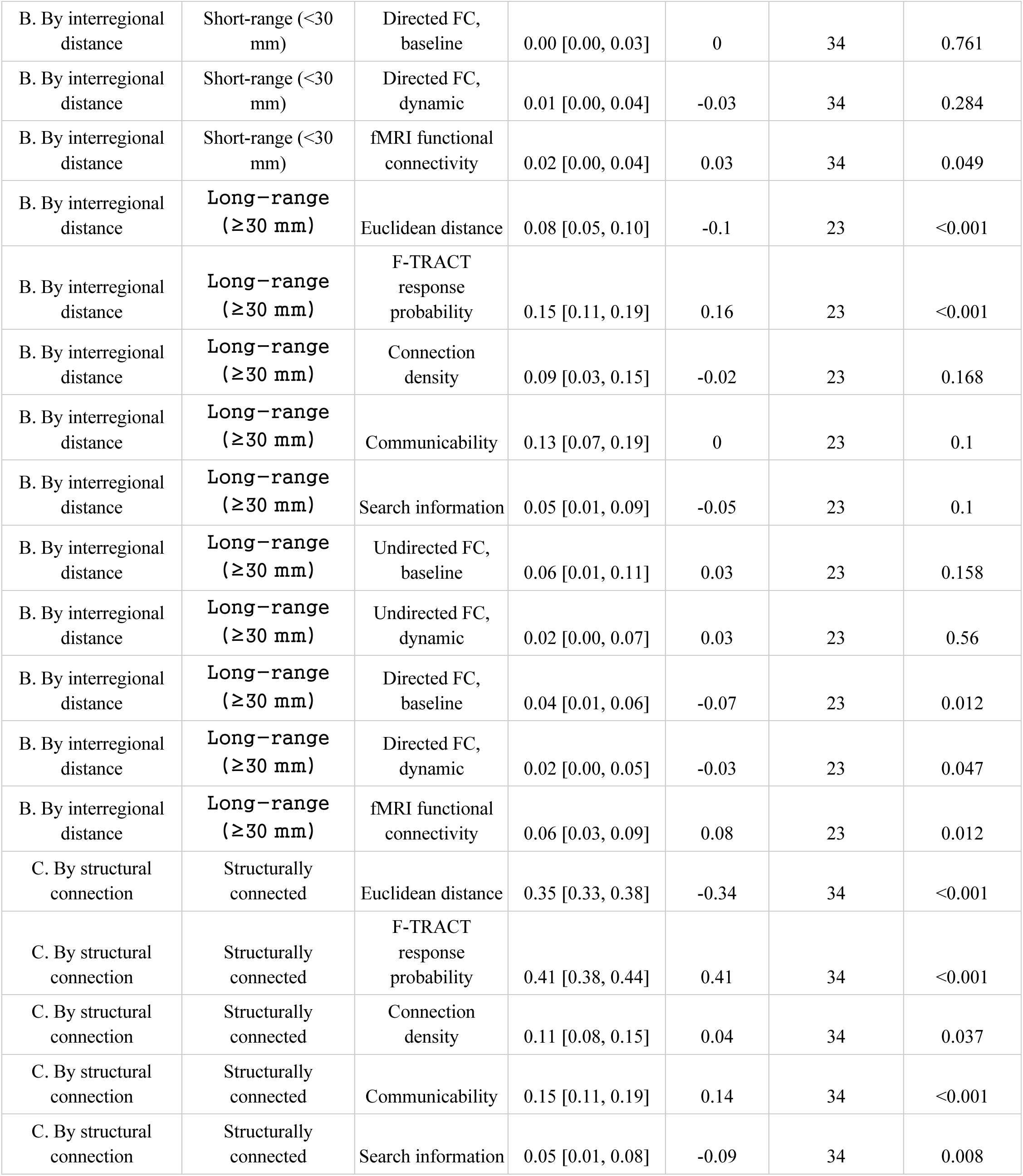
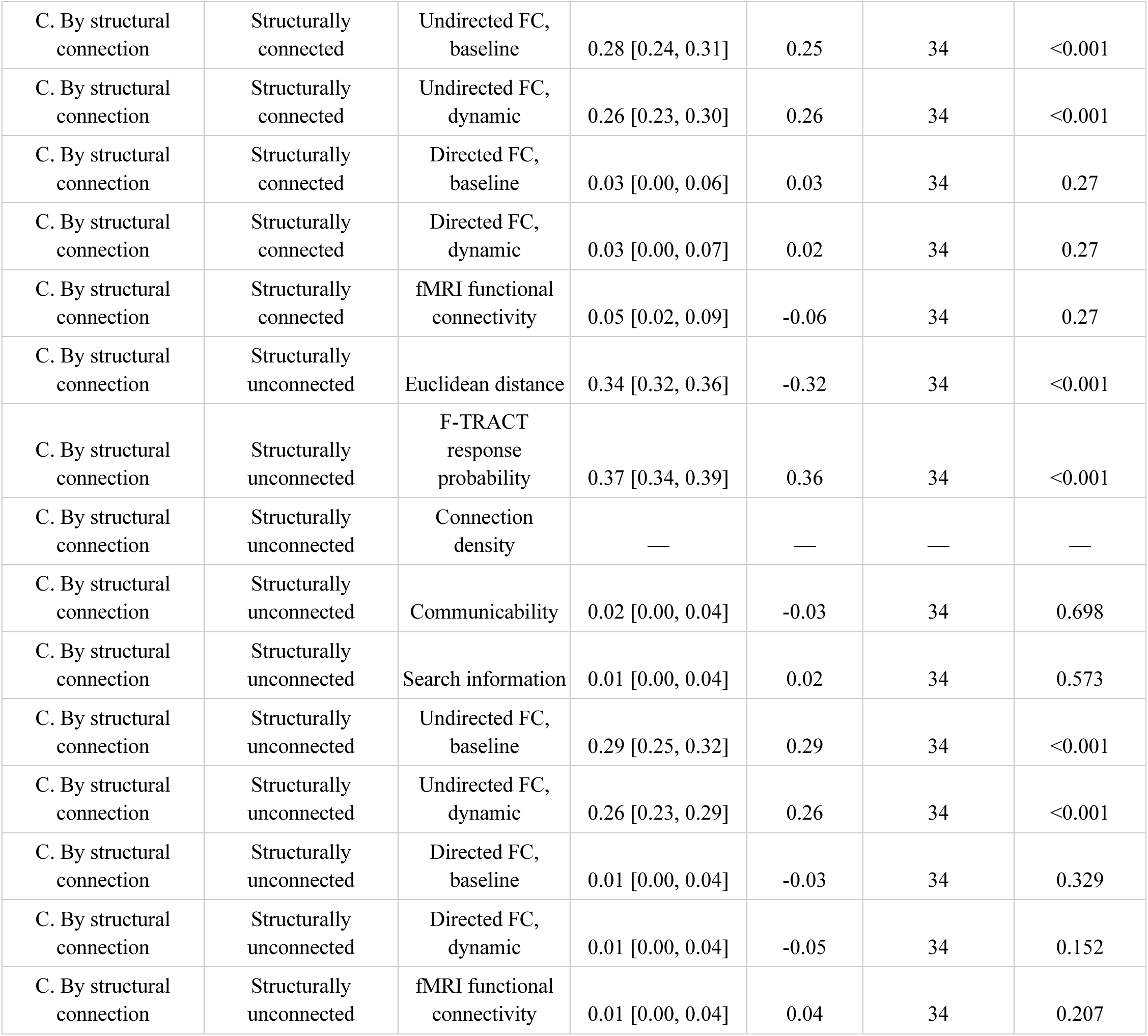
Correlation between features and binary N1 response.

| Panel | Condition | Feature | $\vert \rho\vert $ [95% CI] | Per-patient<br>r, median<br>(signed) | n (patients) | p (FDR) |
| --- | --- | --- | --- | --- | --- | --- |
| A. Control condition<br>(all pairs) | Unadjusted | Euclidean distance | 0.36 [0.35, 0.38] | -0.37 | 34 | <0.001 |
| A. Control condition<br>(all pairs) | Unadjusted | F-TRACT<br>response<br>probability | 0.41 [0.39, 0.43] | 0.42 | 34 | <0.001 |
| A. Control condition<br>(all pairs) | Unadjusted | Connection<br>density | 0.13 [0.10, 0.15] | 0.11 | 34 | <0.001 |
| A. Control condition<br>(all pairs) | Unadjusted | Communicability | 0.16 [0.14, 0.19] | 0.15 | 34 | <0.001 |
| A. Control condition<br>(all pairs) | Unadjusted | Search information | 0.08 [0.06, 0.10] | -0.09 | 34 | <0.001 |
| A. Control condition<br>(all pairs) | Unadjusted | Undirected FC,<br>baseline | 0.29 [0.27, 0.32] | 0.28 | 34 | <0.001 |
| A. Control condition<br>(all pairs) | Unadjusted | Undirected FC,<br>dynamic | 0.27 [0.25, 0.30] | 0.27 | 34 | <0.001 |
| A. Control condition<br>(all pairs) | Unadjusted | Directed FC,<br>baseline | 0.00 [0.00, 0.02] | -0.01 | 34 | 0.919 |
| A. Control condition<br>(all pairs) | Unadjusted | Directed FC,<br>dynamic | 0.02 [0.00, 0.04] | -0.01 | 34 | 0.508 |
| A. Control condition<br>(all pairs) | Unadjusted | fMRI functional<br>connectivity | 0.03 [0.01, 0.05] | 0.06 | 34 | 0.005 |
| A. Control condition<br>(all pairs) | Distance-adjusted | Euclidean distance | 0.36 [0.35, 0.38] | -0.37 | 34 | <0.001 |
| A. Control condition<br>(all pairs) | Distance-adjusted | F-TRACT<br>response<br>probability | 0.23 [0.21, 0.25] | 0.23 | 34 | <0.001 |
| A. Control condition<br>(all pairs) | Distance-adjusted | Connection<br>density | 0.08 [0.05, 0.10] | 0.06 | 34 | 0.022 |
| A. Control condition<br>(all pairs) | Distance-adjusted | Communicability | 0.10 [0.07, 0.12] | 0.07 | 34 | <0.001 |
| A. Control condition<br>(all pairs) | Distance-adjusted | Search information | 0.02 [0.00, 0.04] | -0.04 | 34 | 0.034 |
| A. Control condition<br>(all pairs) | Distance-adjusted | Undirected FC,<br>baseline | 0.17 [0.15, 0.20] | 0.18 | 34 | <0.001 |
| A. Control condition<br>(all pairs) | Distance-adjusted | Undirected FC,<br>dynamic | 0.16 [0.13, 0.18] | 0.16 | 34 | <0.001 |
| A. Control condition<br>(all pairs) | Distance-adjusted | Directed FC,<br>baseline | 0.02 [0.00, 0.04] | -0.02 | 34 | 0.04 |
| A. Control condition (all pairs) | Distance-adjusted | Directed FC, dynamic | 0.00 [0.00, 0.02] | -0.04 | 34 | 0.025 |
| A. Control condition (all pairs) | Distance-adjusted | fMRI functional connectivity | 0.01 [0.00, 0.03] | 0.01 | 34 | 0.134 |
| A. Control condition (all pairs) | F-TRACT-adjusted | Euclidean distance | 0.12 [0.10, 0.14] | -0.12 | 34 | <0.001 |
| A. Control condition (all pairs) | F-TRACT-adjusted | F-TRACT response probability | 0.41 [0.39, 0.43] | 0.42 | 34 | <0.001 |
| A. Control condition (all pairs) | F-TRACT-adjusted | Connection density | 0.02 [0.00, 0.05] | 0 | 34 | 0.904 |
| A. Control condition (all pairs) | F-TRACT-adjusted | Communicability | 0.03 [0.00, 0.05] | 0.01 | 34 | 0.904 |
| A. Control condition (all pairs) | F-TRACT-adjusted | Search information | 0.02 [0.00, 0.04] | 0 | 34 | 0.904 |
| A. Control condition (all pairs) | F-TRACT-adjusted | Undirected FC, baseline | 0.17 [0.14, 0.20] | 0.15 | 34 | <0.001 |
| A. Control condition (all pairs) | F-TRACT-adjusted | Undirected FC, dynamic | 0.15 [0.13, 0.18] | 0.13 | 34 | <0.001 |
| A. Control condition (all pairs) | F-TRACT-adjusted | Directed FC, baseline | 0.01 [0.00, 0.03] | 0 | 34 | 0.946 |
| A. Control condition (all pairs) | F-TRACT-adjusted | Directed FC, dynamic | 0.02 [0.00, 0.04] | -0.02 | 34 | 0.904 |
| A. Control condition (all pairs) | F-TRACT-adjusted | fMRI functional connectivity | 0.01 [0.00, 0.03] | 0 | 34 | 0.904 |
| B. By interregional distance | Short-range (<30 mm) | Euclidean distance | 0.28 [0.26, 0.31] | -0.31 | 34 | <0.001 |
| B. By interregional distance | Short-range (<30 mm) | F-TRACT response probability | 0.33 [0.30, 0.35] | 0.33 | 34 | <0.001 |
| B. By interregional distance | Short-range (<30 mm) | Connection density | 0.11 [0.08, 0.14] | 0.1 | 34 | <0.001 |
| B. By interregional distance | Short-range (<30 mm) | Communicability | 0.14 [0.11, 0.17] | 0.13 | 34 | <0.001 |
| B. By interregional distance | Short-range (<30 mm) | Search information | 0.06 [0.03, 0.09] | -0.08 | 34 | <0.001 |
| B. By interregional distance | Short-range (<30 mm) | Undirected FC, baseline | 0.24 [0.22, 0.27] | 0.23 | 34 | <0.001 |
| B. By interregional distance | Short-range (<30 mm) | Undirected FC, dynamic | 0.23 [0.21, 0.26] | 0.22 | 34 | <0.001 |
| B. By interregional distance | Short-range (<30 mm) | Directed FC, baseline | 0.00 [0.00, 0.03] | 0 | 34 | 0.761 |
| B. By interregional distance | Short-range (<30 mm) | Directed FC, dynamic | 0.01 [0.00, 0.04] | -0.03 | 34 | 0.284 |
| B. By interregional distance | Short-range (<30 mm) | fMRI functional connectivity | 0.02 [0.00, 0.04] | 0.03 | 34 | 0.049 |
| B. By interregional distance | Long-range ($\geq 30$ mm) | Euclidean distance | 0.08 [0.05, 0.10] | -0.1 | 23 | <0.001 |
| B. By interregional distance | Long-range ($\geq 30$ mm) | F-TRACT response probability | 0.15 [0.11, 0.19] | 0.16 | 23 | <0.001 |
| B. By interregional distance | Long-range ($\geq 30$ mm) | Connection density | 0.09 [0.03, 0.15] | -0.02 | 23 | 0.168 |
| B. By interregional distance | Long-range ($\geq 30$ mm) | Communicability | 0.13 [0.07, 0.19] | 0 | 23 | 0.1 |
| B. By interregional distance | Long-range ($\geq 30$ mm) | Search information | 0.05 [0.01, 0.09] | -0.05 | 23 | 0.1 |
| B. By interregional distance | Long-range ($\geq 30$ mm) | Undirected FC, baseline | 0.06 [0.01, 0.11] | 0.03 | 23 | 0.158 |
| B. By interregional distance | Long-range ($\geq 30$ mm) | Undirected FC, dynamic | 0.02 [0.00, 0.07] | 0.03 | 23 | 0.56 |
| B. By interregional distance | Long-range ($\geq 30$ mm) | Directed FC, baseline | 0.04 [0.01, 0.06] | -0.07 | 23 | 0.012 |
| B. By interregional distance | Long-range ($\geq 30$ mm) | Directed FC, dynamic | 0.02 [0.00, 0.05] | -0.03 | 23 | 0.047 |
| B. By interregional distance | Long-range ($\geq 30$ mm) | fMRI functional connectivity | 0.06 [0.03, 0.09] | 0.08 | 23 | 0.012 |
| C. By structural connection | Structurally connected | Euclidean distance | 0.35 [0.33, 0.38] | -0.34 | 34 | <0.001 |
| C. By structural connection | Structurally connected | F-TRACT response probability | 0.41 [0.38, 0.44] | 0.41 | 34 | <0.001 |
| C. By structural connection | Structurally connected | Connection density | 0.11 [0.08, 0.15] | 0.04 | 34 | 0.037 |
| C. By structural connection | Structurally connected | Communicability | 0.15 [0.11, 0.19] | 0.14 | 34 | <0.001 |
| C. By structural connection | Structurally connected | Search information | 0.05 [0.01, 0.08] | -0.09 | 34 | 0.008 |
| C. By structural connection | Structurally connected | Undirected FC, baseline | 0.28 [0.24, 0.31] | 0.25 | 34 | <0.001 |
| C. By structural connection | Structurally connected | Undirected FC, dynamic | 0.26 [0.23, 0.30] | 0.26 | 34 | <0.001 |
| C. By structural connection | Structurally connected | Directed FC, baseline | 0.03 [0.00, 0.06] | 0.03 | 34 | 0.27 |
| C. By structural connection | Structurally connected | Directed FC, dynamic | 0.03 [0.00, 0.07] | 0.02 | 34 | 0.27 |
| C. By structural connection | Structurally connected | fMRI functional connectivity | 0.05 [0.02, 0.09] | -0.06 | 34 | 0.27 |
| C. By structural connection | Structurally unconnected | Euclidean distance | 0.34 [0.32, 0.36] | -0.32 | 34 | <0.001 |
| C. By structural connection | Structurally unconnected | F-TRACT response probability | 0.37 [0.34, 0.39] | 0.36 | 34 | <0.001 |
| C. By structural connection | Structurally unconnected | Connection density | — | — | — | — |
| C. By structural connection | Structurally unconnected | Communicability | 0.02 [0.00, 0.04] | -0.03 | 34 | 0.698 |
| C. By structural connection | Structurally unconnected | Search information | 0.01 [0.00, 0.04] | 0.02 | 34 | 0.573 |
| C. By structural connection | Structurally unconnected | Undirected FC, baseline | 0.29 [0.25, 0.32] | 0.29 | 34 | <0.001 |
| C. By structural connection | Structurally unconnected | Undirected FC, dynamic | 0.26 [0.23, 0.29] | 0.26 | 34 | <0.001 |
| C. By structural connection | Structurally unconnected | Directed FC, baseline | 0.01 [0.00, 0.04] | -0.03 | 34 | 0.329 |
| C. By structural connection | Structurally unconnected | Directed FC, dynamic | 0.01 [0.00, 0.04] | -0.05 | 34 | 0.152 |
| C. By structural connection | Structurally unconnected | fMRI functional connectivity | 0.01 [0.00, 0.04] | 0.04 | 34 | 0.207 |

**Table S2a:** Model performance (AUC)

| Model | n (patients) | AUC, median [IQR] | AUPRC, median [IQR] | AUC, mean ± SD |
| --- | --- | --- | --- | --- |
| Euclidean distance (ED) | 34 | 0.803 [0.761, 0.847] | 0.442 [0.396, 0.532] | 0.801 ± 0.059 |
| ED + F-TRACT | 34 | 0.825 [0.786, 0.861] | 0.502 [0.418, 0.580] | 0.821 ± 0.055 |
| ED + F-TRACT + structural | 34 | 0.828 [0.787, 0.864] | 0.504 [0.400, 0.593] | 0.820 ± 0.054 |
| ED + F-TRACT + structural + fMRI | 34 | 0.827 [0.788, 0.865] | 0.500 [0.401, 0.595] | 0.821 ± 0.054 |
| Full model (+ iEEG FC) | 34 | 0.840 [0.794, 0.875] | 0.523 [0.431, 0.587] | 0.833 ± 0.049 |

**Table S2b:** Incremental-model comparisons.

| Comparison | $\Delta$ AUC (median) | p (FDR) | Significant |
| --- | --- | --- | --- |
| Euclidean distance (ED) vs ED + F-TRACT | 0.022 | <0.001 | Yes |
| ED + F-TRACT vs ED + F-TRACT + structural | 0.001 | 0.853 | No |
| ED + F-TRACT + structural vs ED + F-TRACT + structural + fMRI | 0 | 0.853 | No |
| ED + F-TRACT + structural + fMRI vs Full model (+ iEEG FC) | 0.012 | <0.001 | Yes |

**Table S2c:** Predicted response probabilities.

| True class | n (edges) | Predicted P(response), median [IQR] |
| --- | --- | --- |
| No Response | 7548 | 0.27 [0.13, 0.52] |
| Response | 1635 | 0.69 [0.52, 0.84] |

**Table S3:**
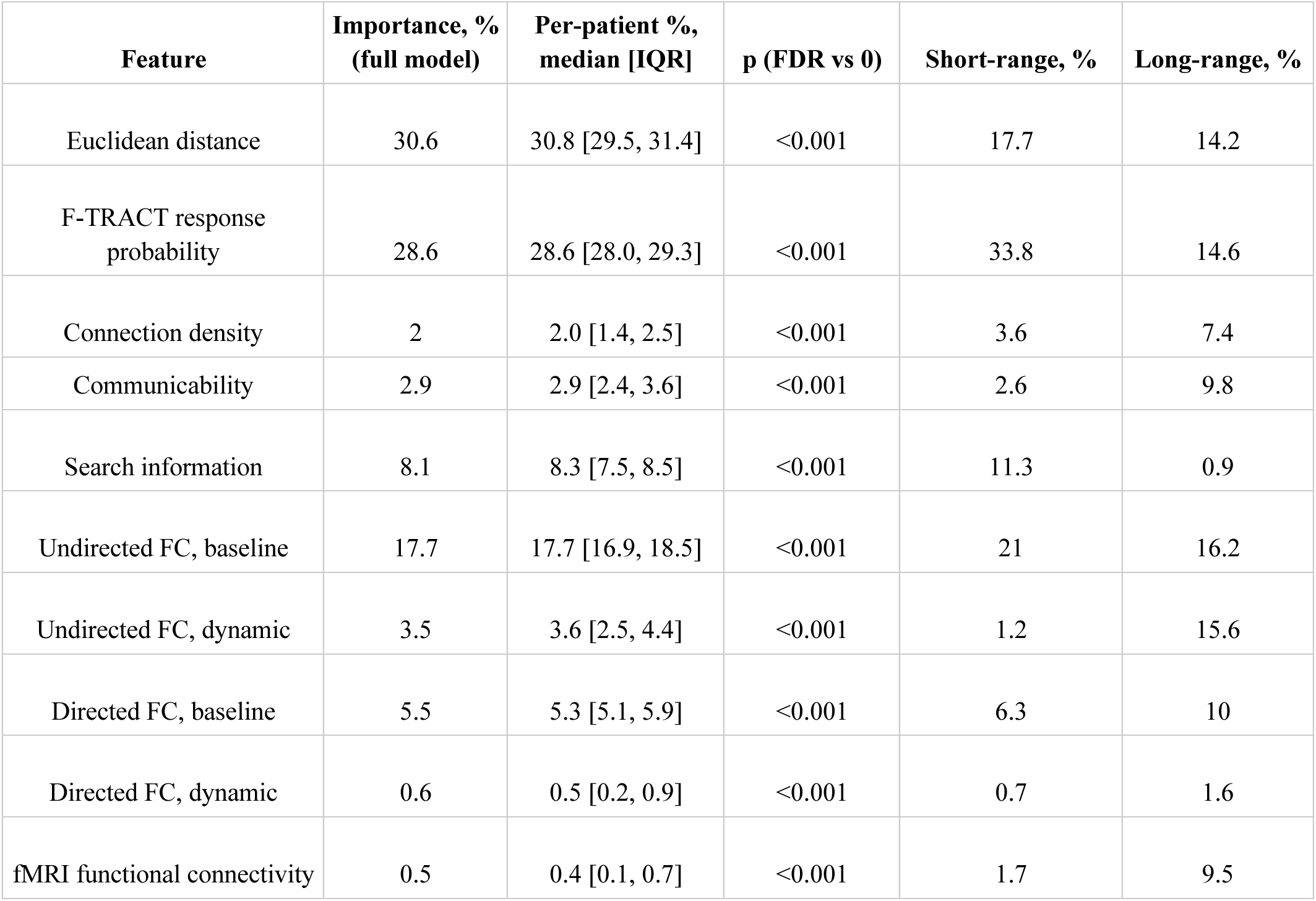
Feature importance for full model.

**Table S4a:**
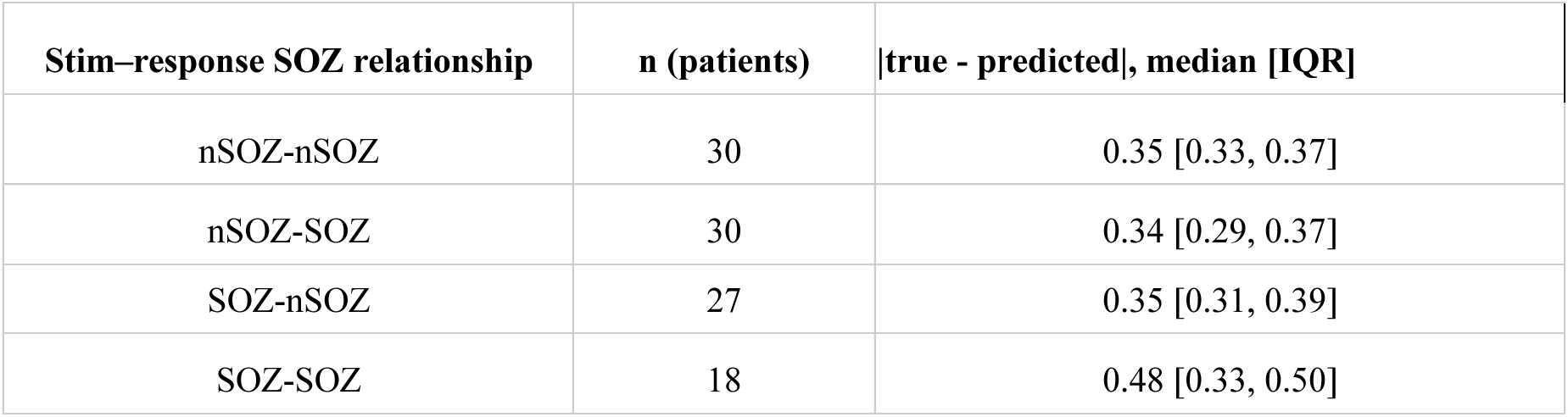
Prediction residuals by SOZ relationship.

| **Stim-response SOZ relationship** | **n (patients)** | **\|true - predicted\|, median [IQR]** |
| --- | --- | --- |
| nSOZ-nSOZ | 30 | 0.35 [0.33, 0.37] |
| nSOZ-SOZ | 30 | 0.34 [0.29, 0.37] |
| SOZ-nSOZ | 27 | 0.35 [0.31, 0.39] |
| SOZ-SOZ | 18 | 0.48 [0.33, 0.50] |

**Table S4b:**
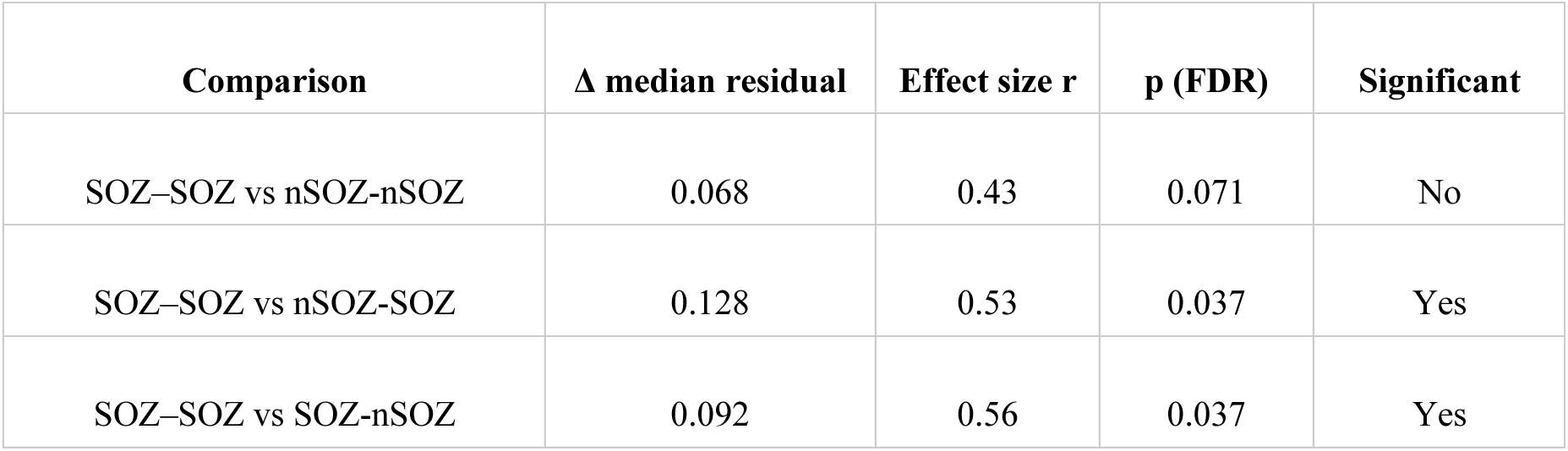
Residual comparisons (SOZ–SOZ vs each)

**Table S4c:** Error rates by SOZ relationship.

| Stim–response SOZ relationship | FPR, pair-level [95% CI] | FNR, pair-level [95% CI] | FPR, patient median | FNR, patient median |
| --- | --- | --- | --- | --- |
| nSOZ-nSOZ | 0.28 [0.26, 0.29] | 0.24 [0.21, 0.26] | 0.28 | 0.21 |
| nSOZ-SOZ | 0.27 [0.24, 0.30] | 0.12 [0.08, 0.19] | 0.29 | 0.04 |
| SOZ-nSOZ | 0.29 [0.26, 0.33] | 0.28 [0.21, 0.36] | 0.29 | 0.22 |
| SOZ-SOZ | 0.53 [0.41, 0.65] | 0.00 [0.00, 0.08] | 1 | 0 |

**Table S4d:** Error-rate comparisons (SOZ–SOZ vs each)

| Comparison | Metric | Effect size r | p (FDR) | Significant |
| --- | --- | --- | --- | --- |
| SOZ–SOZ vs nSOZ-nSOZ | FPR | 0.87 | 0.001 | Yes |
| SOZ–SOZ vs nSOZ-SOZ | FPR | 0.87 | 0.001 | Yes |
| SOZ–SOZ vs SOZ-nSOZ | FPR | 0.87 | 0.001 | Yes |
| SOZ–SOZ vs nSOZ–nSOZ | FNR | 0.88 | 0.002 | Yes |
| SOZ–SOZ vs nSOZ–SOZ | FNR | 0.99 | 0.009 | Yes |
| SOZ–SOZ vs SOZ–nSOZ | FNR | 0.96 | 0.004 | Yes |

**Table S5a:**
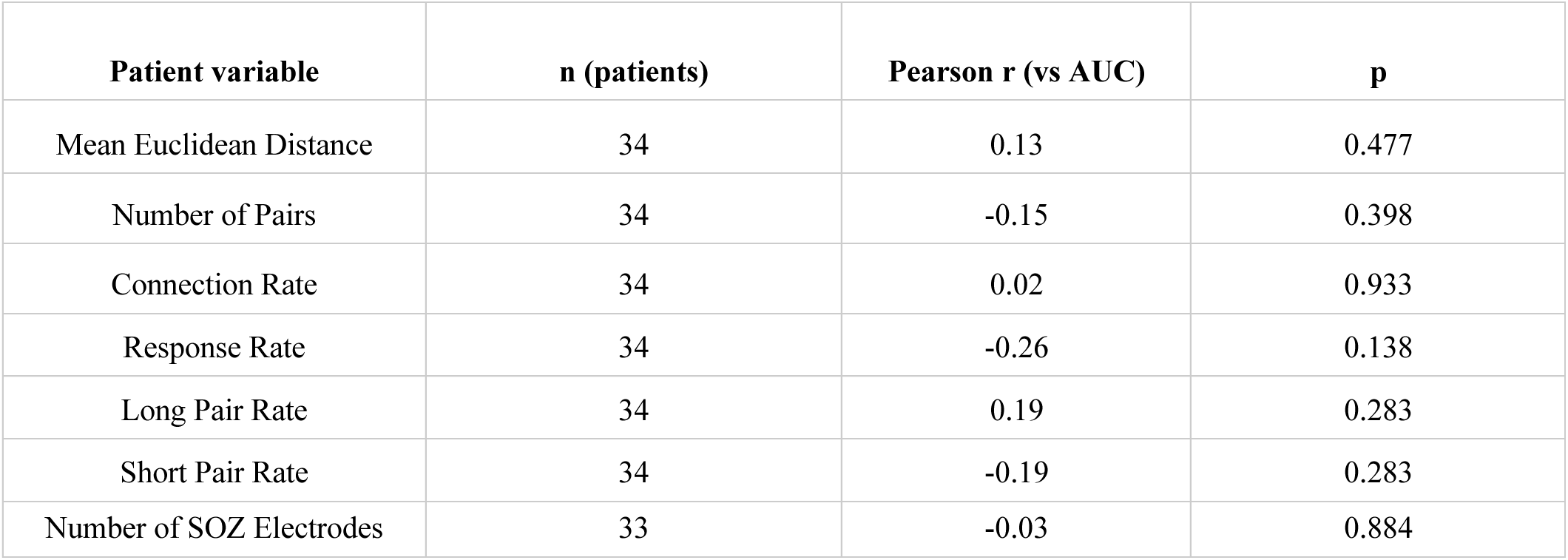
Continuous clinical/implant confounders.

**Table S5b:** Categorical clinical confounders (AUC)

| Variable | Category | n | AUC, median [IQR] | Pairwise test (Bonferroni) |
| --- | --- | --- | --- | --- |
| Intervention | DBS | 1 | 0.839 [0.839, 0.839] | all n.s. |
|  | Laser Ablation | 11 | 0.841 [0.808, 0.881] |  |
|  | No intervention | 9 | 0.805 [0.789, 0.840] |  |
|  | RNS | 3 | 0.881 [0.839, 0.886] |  |
|  | Resection | 9 | 0.864 [0.834, 0.870] |  |
|  | VNS | 1 | 0.784 [0.784, 0.784] |  |
| Implant Lateralization | Bilateral | 8 | 0.812 [0.796, 0.840] | all n.s. |
|  | Left > Right | 3 | 0.762 [0.744, 0.822] |  |
|  | Right > Left | 3 | 0.840 [0.814, 0.858] |  |
|  | Unilateral | 18 | 0.860 [0.825, 0.888] |  |
| Imaging Findings | Nonspecific Abnormality | 12 | 0.840 [0.790, 0.867] | all n.s. |
|  | Normal | 9 | 0.844 [0.823, 0.872] |  |
|  | Potential Epileptogenic Lesion | 12 | 0.834 [0.783, 0.884] |  |
| Sex | Female | 19 | 0.840 [0.795, 0.873] | all n.s. |
|  | Male | 15 | 0.833 [0.793, 0.877] |  |

**Table S5c:** AUC by SOZ lateralization / region.

| Variable | Category | n | AUC, median [IQR] | Pairwise test (Bonferroni) |
| --- | --- | --- | --- | --- |
| SOZ Region | Mesial temporal | 8 | 0.808 [0.790, 0.873] | all n.s. |
|  | Multifocal | 4 | 0.840 [0.825, 0.850] |  |
|  | Other cortex | 1 | 0.896 [0.896, 0.896] |  |
|  | Temporal | 5 | 0.833 [0.762, 0.854] |  |
|  | Temporal neocortical | 3 | 0.834 [0.780, 0.853] |  |
| SOZ Lateralization | Bilateral | 6 | 0.822 [0.796, 0.840] | all n.s. |
|  | Left | 16 | 0.826 [0.776, 0.856] |  |
|  | Right | 8 | 0.871 [0.856, 0.885] |  |

**Table S5d:** SOZ electrode count by laterality.

| Variable | Category | n | SOZ electrodes, median [IQR] |
| --- | --- | --- | --- |
| SOZ Lateralization | Bilateral | 6 | 12.5 [12.0, 17.5] |
|  | Left | 16 | 5.5 [3.8, 8.2] |
|  | Right | 8 | 7.0 [5.8, 7.2] |
| Implant Lateralization | Bilateral | 7 | 7.0 [3.0, 11.0] |
|  | Left > Right | 3 | 4.0 [3.5, 6.0] |
|  | Right > Left | 3 | 19.0 [16.0, 19.5] |
|  | Unilateral | 18 | 6.0 [4.2, 7.0] |

